# Evaluation of accuracy, exclusivity, limit-of-detection and ease-of-use of LumiraDx™-Antigen-detecting point-of-care device for *SARS-CoV-2*

**DOI:** 10.1101/2021.03.02.21252430

**Authors:** Lisa J. Krüger, Julian A.F. Klein, Frank Tobian, Mary Gaeddert, Federica Lainati, Sarah Klemm, Paul Schnitzler, Ralf Bartenschlager, Berati Cerikan, Christopher J. Neufeldt, Olga Nikolai, Andreas K. Lindner, Frank P. Mockenhaupt, Joachim Seybold, Terry C. Jones, Victor M. Corman, Nira R. Pollock, Britta Knorr, Andreas Welker, Margaretha de Vos, Jilian A. Sacks, Claudia M. Denkinger, for the study team

## Abstract

**Background:** Rapid antigen-detecting tests (Ag-RDTs) for severe acute respiratory syndrome coronavirus 2 (*SARS-CoV-2*) can transform pandemic control. Thus far, sensitivity (≤85%) of lateral-flow assays has limited scale-up. Conceivably, microfluidic immunofluorescence Ag-RDTs could increase sensitivity for *SARS-CoV-2* detection.

**Materials and Methods:** This multi-centre diagnostic accuracy study investigated performance of the microfluidic immunofluorescence LumiraDx™ assay, enrolling symptomatic and asymptomatic participants with suspected *SARS-CoV-2* infection. Participants collected a supervised nasal mid-turbinate (NMT) self-swab for Ag-RDT testing, in addition to a professionally-collected nasopharyngeal (NP) swab for routine testing with reverse transcriptase polymerase chain reaction (RT-PCR). Results were compared to calculate sensitivity and specificity. Sub-analyses investigated the results by viral load, symptom presence and duration. An analytical study assessed exclusivity and limit-of-detection (LOD). In addition, we evaluated ease-of-use.

**Results:** Study conduct was between November 2^nd^ 2020 and January 21^st^ 2021. 761 participants were enrolled, with 486 participants reporting symptoms on testing day. 120 out of 146 RT-PCR positive cases were detected positive by LumiraDx™, resulting in a sensitivity of 82.2% (95% CI: 75.2%-87.5%). Specificity was 99.3% (CI: 98.3-99.7%). Sensitivity was increased in individuals with viral load ≥ 7 log10 SARS-CoV2 RNA copies/ml (93.8%; CI: 86.2%-97.3%). Testing against common respiratory commensals and pathogens showed no cross-reactivity and LOD was estimated to be 2-56 PFU/mL. The ease-of-use-assessment was favourable for lower throughput settings.

**Conclusion:** The LumiraDx™ assay showed excellent analytical sensitivity, exclusivity and clinical specificity with good clinical sensitivity using supervised NMT self-sampling.

## Introduction

The *SARS-CoV-2* pandemic has caused over 100 million confirmed infections worldwide, stated by the World Health Organization (WHO) (https://covid19.who.int/). Reverse transcriptase polymerase chain reaction (RT-PCR) has been established as the gold standard for diagnosing individuals infected with *SARS-CoV-2* (1). Because RT-PCR testing requires advanced laboratory infrastructure demanding numerous predefined materials and specially-trained staff, testing capacities in many countries have at times been pushed to their limits. Also, the longer turn-around time for results from laboratory-based RT-PCR testing does not allow for rapid decision-making, often taking days until results are available, thus identifying infected individuals often too late to prevent secondary cases, expressed by the WHO (https://www.who.int/dg/speeches/detail/who-director-general-s-opening-remarks-at-the-media-briefing-on-covid-19---16-march-2020).

*SARS-CoV-2* rapid antigen-detecting tests (Ag-RDTs) can be considered complementary to RT-PCR for rapid testing, or as an alternative for RT-PCR in settings where resources and laboratory infrastructure are limited as suggested by the WHO (https://www.who.int/news-room/commentaries/detail/advice-on-the-use-of-point-of-care-immunodiagnostic-tests-for-covid-19). Several countries are incorporating these tests into their national testing strategies and some (e.g. Slovakia and the United Kingdom) are utilizing them for large scale screening (2).

Most tests on the market use the lateral flow principle, gathered and explained by the Foundation of Innovative New Diagnostics (FIND) (https://www.finddx.org/covid-19/pipeline/). While many tests have shown excellent specificity, sensitivity in several independent evaluations has been around 85% or less (https://diagnosticsglobalhealth.org/ (3–5). Conceivably, microfluidic immunofluorescence combined with digital result capture and connectivity could boost sensitivity and ease of use of Ag-RDTs (6).

LumiraDx™ has recently developed a diagnostic device using microfluidic immunofluorescence technology with automated read-out to enable rapid detection of *SARS-CoV-2* antigen from a nasal mid-turbinate (NMT) swab (https://www.lumiradx.com/uk-en/what-we-do/diagnostics/test-technology/antigen-test). In the United States, the LumiraDx™ *SARS-CoV-2* Ag-RDT was granted an emergency use authorization by the Federal Drug Agency (FDA) in August 2020 (https://www.fda.gov/emergency-preparedness-and-response/mcm-legal-regulatory-and-policy-framework/emergency-use-authorization). In the United Kingdom, a large pharmacy chain offers LumiraDx™ *SARS-CoV-2* testing at the point-of-care (https://www.boots-uk.com/media-centre/press-releases/new-covid-19-testing-service-now-available-in-boots-uk-stores/). To date, there is only one publication available, sponsored by the manufacturer, that reported a 97.6% sensitivity and 96.6% specificity (7) in nasal swabs from mostly symptomatic persons within 12 days of symptom onset.

This multi-centre, manufacturer-independent diagnostic accuracy study investigated the performance, ease-of-use, exclusivity, and limit-of-detection of the microfluidic immunofluorescence *SARS-CoV-2* LumiraDx™ antigen test (henceforth called LumiraDx™).

## Materials and Methods

### Index Test

The diagnostic test under evaluation in this study was the microfluidic immunofluorescence assay, *SARS-CoV-2* Ag test developed by LumiraDx™ Limited, London, United Kingdom (henceforth called LumiraDx™). The assay runs on a portable platform using a dry, single-use, disposable, microfluidic test strip. The strip incorporates antibodies specific to the nucleocapsid protein of *SARS-CoV-2*, that, in combination with magnetic particles, form a sandwich-like immuno-complex that emits a fluorescent latex signal (https://www.lumiradx.com/uk-en/what-we-do/diagnostics/test-technology/antigen-test).

The sample collection was performed with Dryswab™ Standard Tip Rayon (Medical Wire & Equipment, Corsham, England), not included in the test kits, but recommend for use by the manufacturer. Ag-RDT testing was performed on-site in a dedicated workspace, that was divided into separate areas where infectious and non-infectious materials were handled. Laboratory personal for Ag-RDT testing was blinded to the results of RT-PCR testing and vice versa. Following manufacturer instructions for use, the collected sample was processed using the LumiraDx™ proprietary extraction buffer vial for ten seconds. A single-use test strip was placed in the designated slot on the testing device, and one drop (∼ 20 μL) of the prepared extraction buffer solution was applied. The device was closed to initiate automatic processing and testing. The device showed a qualitative result (“positive”, “negative” or “error”) on the digital touch screen within twelve minutes. In case of an erroneous test result, the testing was immediately repeated using the solution from the same extraction vial and a new test strip.

For every test, a cut-off-index (COI) value was automatically generated and documented in the testing device, reporting the test immunofluorescent signal on a continuous scale with a pre-set cut-off categorizing the results as positive or negative. COI values were retrieved from the test devices by the manufacturer after completion of the study. The device can operate in a cloud-based mode enabling the manufacturer to receive all impersonal data through an online connection. During this study, the manufacturer could not access the device as the study was performed offline.

After every test, the surface of the device was disinfected with proprietary disinfection wipes provided by the company. A five-minute drying time was recommended before the next sample was inserted, following the manufacturer information for use (https://www.lumiradx.com/uk-en/what-we-do/diagnostics/test-technology/antigen-test).

## Reference test

Reverse transcriptase-polymerase chain reaction (RT-PCR) was used as the reference test. The RT-PCR samples were collected by health-care workers using the IMPROSWAB® (Guangzhou Improve Medical Instruments Co., Ltd., Guangzhou, China) at the Heidelberg study site and the eSwab™ (Copan Diagnostics Inc., Murrieta, CA, USA) at the study site in Berlin. Collected samples were transferred to the referral laboratories for RT-PCR testing in Heidelberg and Berlin. The RT-PCR assays, referral laboratory standard assays, used for clinical diagnosis as comparators were Allplex™ *SARS-CoV-2* assay (Seegene, Seoul, South Korea) in Heidelberg and cobas™ SARS CoV-2 assay on the cobas® 6800 or 8800 system (Roche, Pleasanton, CA, United States) or the SARS CoV-2 assay from TIB Molbiol (Berlin, Germany) in Berlin. Interpretation of the RT-PCR assays followed the manufacturers’ instructions. The RT-PCR assays targeted the E gene of SARS-CoV-2, which was used for the CT-value determination and the viral load calculations. Based on testing of standardized material, CT-values of the three tests are expected to differ by around 2-3 with the same amount of virus present [2]. The conversion of CT-values into viral-load was based on calibrated RT-PCR testing with quantified *SARS-CoV-2* in vitro transcripts [2].

### Clinical diagnostic accuracy

The standards for reporting diagnostic accuracy studies (STARD) were followed for reporting of this study (8).

#### Study design and inclusion criteria

This prospective multi-centre study enrolled participants at two sites, Heidelberg and Berlin, Germany. In Heidelberg, participants were enrolled at a drive-in testing site whereas in Berlin participants were enrolled at a clinical ambulatory testing facility. All participants were identified for testing according to the criteria of the national health authority as being at risk of *SARS-CoV-2* infection based on reported symptoms or recent contact with a confirmed case (https://www.rki.de/DE/Content/InfAZ/N/Neuartiges_Coronavirus/Teststrategie/Testkriterien_Herbst_Winter.html). Inclusion criteria were a minimum age of 18 years and no prior positive RT-PCR result for *SARS-CoV-2*. Written informed consent was obtained from each participant prior to testing. Individuals unable to give written informed consent due to limited command of German or English were excluded from the study. The study protocol is available upon request.

#### Sample collection

The sample for the Ag-RDT was a NMT, following the definition from the Centers for Disease Control and Prevention (https://www.cdc.gov/coronavirus/2019-ncov/lab/guidelines-clinical-specimens.html#specimen.). This swab was collected by the participants themselves, with a health-care worker providing instructions, supervision and corrections according to the instruction for use by the manufacturer. The participants were instructed to tilt their head back 70 degrees, insert the swab approximately 2cm into one nostril, and to rotate it several times against the full interior nasal wall surface for at least 10 seconds. The identical procedure was repeated with the same swab in the contralateral nostril.

Subsequently, the professional-collected routine swab for RT-PCR testing was taken, following institutional procedures with a nasopharyngeal (NP) swab (Heidelberg) or a combined NP/oropharyngeal (OP) swab (Berlin). Participants underwent OP swabbing only if there were clinical contraindications for NP sampling.

#### Questionnaire

All participants were asked for information on comorbidities, symptom presence and duration, and severity of disease (questionnaire available in the supplement material, Section (A)). For data collection, we used the Research Electronic Data Capture (REDCap) tools hosted at Heidelberg University (9).

#### RT-PCR from Ag-RDT extraction buffer

All false-positive and false-negative Ag-RDT results were retested with RT-PCR from the Ag-RDT proprietary buffer solution that was stored at a temperature of −20°C if sufficient volume was available. This testing is not validated by the manufacturer but was performed to resolve discrepant results that could have occurred due to variability between the NP and NMT sampling for RT-PCR and Ag-RDT. To avoid the introduction of a discrepant analysis bias, a subset of samples (first 5 per day in the first 2 weeks) was also re-tested with RT-PCR from the antigen buffer.

### Exclusivity testing

Clinical samples from common respiratory tract pathogens or commensals were assessed for cross-reactivity. The respiratory swab samples contained coronavirus, adenovirus, bocavirus, influenza virus, metapneumovirus, parainfluenza virus, respiratory syncytial virus, rhinovirus or *Mycoplasma pneumoniae* as identified by RT-PCR in the laboratory. For *Staphylococcus aureus* and *Streptococcus sp*. swab samples from bacterial culture were utilized. As recommended by the manufacturer these samples were added to the extraction buffer in a 1:10 dilution (30µl sample and 270µl extraction buffer) and processed according to the standard test protocol.

### Limit-of-detection assay using cell culture-grown SARS-CoV-2

The BavPat1/2020 strain was kindly provided by Christian Drosten through the European Virus Archive and the HD strain was isolated from a patient at the Heidelberg clinic. Working virus stocks were generated by passaging the virus two times in VeroE6 cells or Calu3 cells (a kind gift from Dr. Manfred Frey, Mannheim). Virus stocks were tittered using a plaque forming unit (PFU) assay (10). The stock was pre-diluted in DMEM supplemented with 2% FCS to 10000 PFU/mL. Then two-fold dilutions were generated and added 1:1 to the isolation buffer provided for each test kit. Following this, the manufacturer’s protocol for each kit was used. Note, that the amount of volume used for each kit was slightly different according to the protocols (50µL for Abbott PanBio, 100µL for Biosensor Standard Q and 35µL for LumiraDx™). The limit of detection was calculated based on the number of PFUs in this volume. Three replicates were done for each dilution.

### System Usability Scale and Ease-of-Use Assessment

To determine and quantify the usability of the test, a standardized System Usability Scale (SUS) questionnaire was used and a dedicated ease-of-use assessment (EoU) was developed. The detailed surveys are provided in the supplement material (Section (B) and (C)). Staff performing the testing at both study sites were invited to complete the questionnaires. A SUS score above 68 is interpreted as above average (11). For the visualization of the EoU assessment a colour-coded rating (heat-map) was generated. Each aspect of the assessment was ranked as satisfactory, average, or unsatisfactory. The supplement material (Section (D)) shows the matrix used for this analysis.

### Statistics

To determine sensitivity and specificity of the Ag-RDT (with 95% CIs), results were compared to RT-PCR results from the same participant, as per Altman (12). Predefined sub-analyses were conducted for presence of symptoms, duration of symptoms (≤7, >7 days and >12 days), CT-values (using two categorizations: ≤25, >25 and ≤30, >30), viral load, and study site. A two-sided alpha value of 0.05 was defined as a significance cut-off. We used “R” version 4.0.3. (R Foundation for Statistical Computing, Vienna, Austria) to generate all analyses and plots.

### Study approval

The study protocol was approved in March 2020 by the ethical review committee at the Heidelberg University Hospital for the two study sites Heidelberg and Berlin in Germany (Registration number S-180/2020). All participants provided written informed consent prior to inclusion in the study. The study was registered in the German clinical trial registry with the registration number DRKS00021220.

## Results

A total of 826 participants were screened for enrolment between the 2^nd^ November and 4^th^ December 2020 across two study sites, and 767 (92.9%) participants gave written informed consent. Of the 767 participants, 493 were enrolled in Heidelberg and 274 in Berlin. Two participants were excluded after enrolment as the NMT swab was refused and four participants were excluded during analysis due to invalid RT-PCR results, resulting in 761 participants included in the analysis (Figure 1).

**Figure 1.**
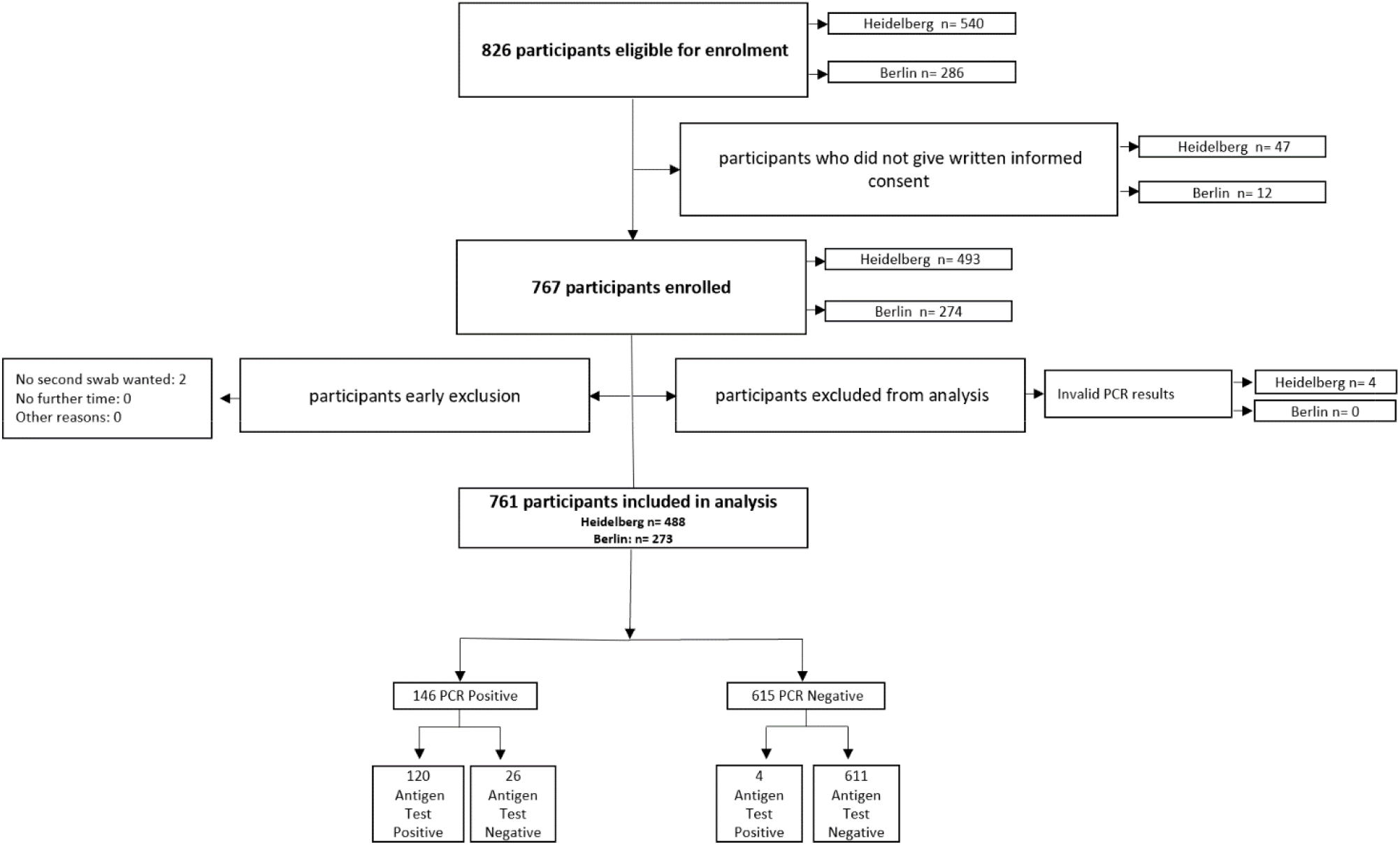
Study Flow.

Table 1 shows the clinical and demographic characteristics of all enrolled participants. The mean age of the participants was 38.5 years (SD, 14.2) with 52% female and 31.7% with comorbidities (Table 1). Symptoms on the testing day were reported by 64% of participants, with an average symptom duration of 3.9 days (SD 3.2). Supplementary a detailed overview with all reported symptoms is found in Section (E) Table 1. All asymptomatic participants were recent high-risk contacts. From a total of 472 symptomatic patients, 423 (89.6%) reported onset of symptoms within the prior 7 days. A total of 146 (19.2%) participants had a positive *SARS-CoV-2* RT-PCR result, leading to a positivity rate of 29.7% (81 participants) in Berlin and 13.3% (65 participants) in Heidelberg. Of all symptomatic participants a total of 137 (28.2%) had a positive *SARS-CoV-2* RT-PCR result whereas only 9 (3.3%) of the asymptomatic participants had a *SARS-CoV-2* RT-PCR result. The median viral load was 7.26 log10 SARS-CoV2 RNA copies/ml, with Berlin and Heidelberg differing only slightly (7.75 and 6.97, respectively). Seven tests resulted in an error message by the LumiraDx™ device. Upon repeat from the same sample extraction buffer, all samples yielded valid results and were included in the analysis. Almost all participants were able to perform the self-sampling without issues, with only 8 participants (1.1%) minorly deviating from the sample collection protocol.

**Table 1.** Study population characteristics.

|  |  | **Overall**<br>**Total N=**<br>**761** | **Heidelberg**<br>**Total N=**<br>**488** | **Berlin**<br>**Total N=**<br>**273** |
| --- | --- | --- | --- | --- |
| **Age in years**<br>Information available on N=761 | Mean<br>(SD) | 38.5<br>(14.2) | 40.1<br>(14.9) | 35.8<br>(12.3) |
| **Gender**<br>Information available on N=759 | Female<br>N (%) | 396<br>(52.2) | 266<br>(54.7) | 130<br>(47.6) |
| **BMI >25**<br>Information available on<br>N=703 | N<br>(%) | 327<br>(46.5) | 243<br>(50.2) | 84<br>(38.4) |
| **Comorbidities**<br>Information available on<br>N=760 | N<br>(%) | 241<br>(31.7) | 183<br>(37.6) | 58<br>(21.2) |
| **Symptoms on Testing Day**<br>Information available on N= 758 | N<br>(%) | 486<br>(64.1) | 218<br>(44.8) | 268<br>(98.2) |
| **Duration of symptoms from day of**<br>**testing in days**<br>Information available on N= 472 | Mean<br>(SD) | 3.9<br>(3.2) | 3.7<br>(3.7) | 4.1<br>(2.7) |
| **Prior test performed and negative**<br>Information available on N= 695 | Yes<br>N (%) | 86<br>(12.4) | 80<br>(16.6) | 6<br>(2.8) |
| **RT-PCR positives** | N<br>(%) | 146<br>(19.2) | 65<br>(13.3) | 81<br>(29.7) |
| **Viral Load**<br>(log10 SARS-CoV2 RNA copies/ml)<br>Information available on N= 146 | Median<br>(IQR) | 7.3<br>(3.7-9.4) | 7.0<br>(3.4-8.5) | 7.8<br>(3.8-9.4) |

LumiraDx™ had an overall sensitivity of 82.2% (120 of 146 RT-PCR positives detected; 95% CI: 75.2%-87.5%) and a specificity of 99.3% (95% CI: 98.3-99.7%) (Figure 2). The performance varied only slightly by site, with sensitivity of 84.6% (95% CI: 7.9%-91.4%) and 80.2% (95% CI: 70.3-87.5%) and specificity of 99.3% (95% CI: 97.9-99.7%) and 99.5% (95% CI: 97.1%-100%) in Heidelberg and Berlin, respectively. The sensitivity was 86.9% (95% CI: 83.0%-93.8%) for samples with a viral load ≥ 5 log10 SARS-CoV2 RNA copies/ml and 93.8% (95% CI: 86.2%-97.3%) for samples with a viral load ≥ 7 log10 SARS-CoV2 RNA copies/ml (Figure 2). Sensitivity in different categories of viral load compared to the RT-PCR results is shown in Figure 3. Similar, for samples with cycle threshold (CT) values <25, the sensitivity was 92.6% (95% CI: 85.6%-96.4%) and for those with a CT-value <30, the sensitivity was 90.2% (95% CI: 83.6%-94.3%) (Figure 2). Correspondingly, the sensitivity was 62.7% (95% CI: 49.0%-74.7%) and 41.7% (95% CI: 24.5%-61.2%) for samples with a CT-value ≥ 25 and CT-value ≥ 30 (Figure 2). Supplementary Table 2 (Section (F)) shows a detailed listing of the viral load, CT-value and Ag-RDT result for each participant. Overall a moderate correlation was observed between the signal intensity (measured by the cut-off-index (COI) values) of the LumiraDx™ device and viral load, except for a few high viral load samples (5 with viral load >7 log10 SARS-CoV2 RNA copies/ml) that scored a low COI and were considered false-negative (supplement Section (G) Figure 2). Spearman rank correlation coefficient was calculated to be 0.57 between COI and viral load.

**Figure 2.**
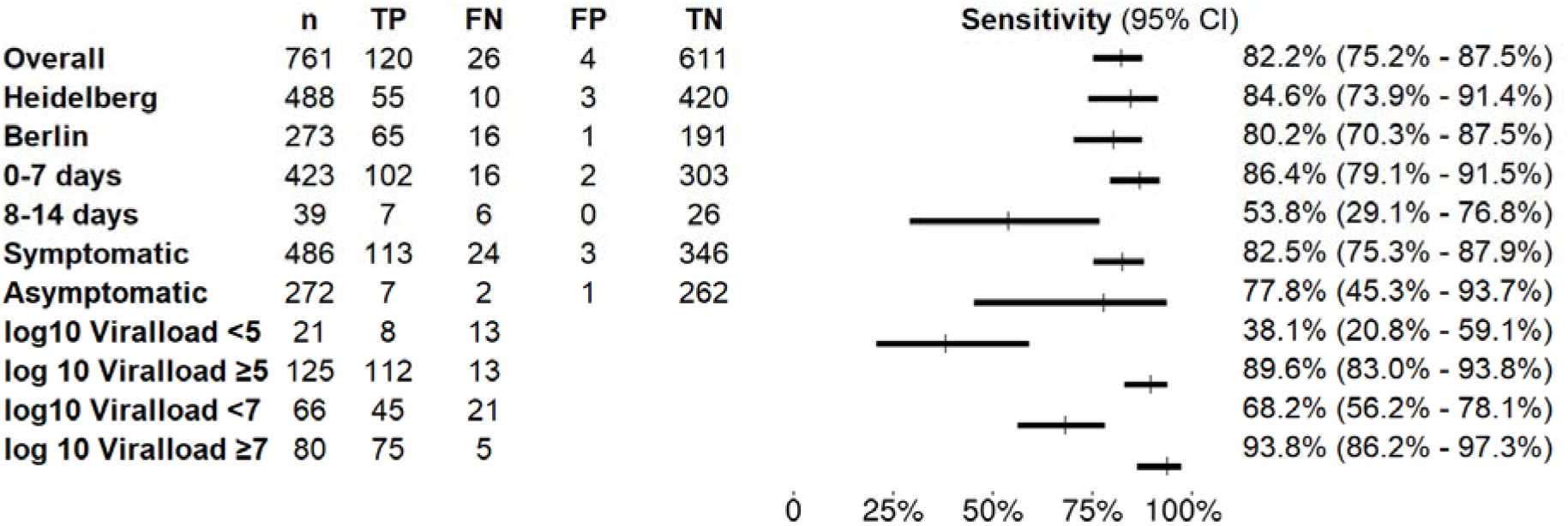
Forest plot of sensitivity analysis overall and by subgroup for LumiraDx™. n = number, TP = true positives, FN = false negatives, FP = false positives, TN = true negatives

**Figure 3.**
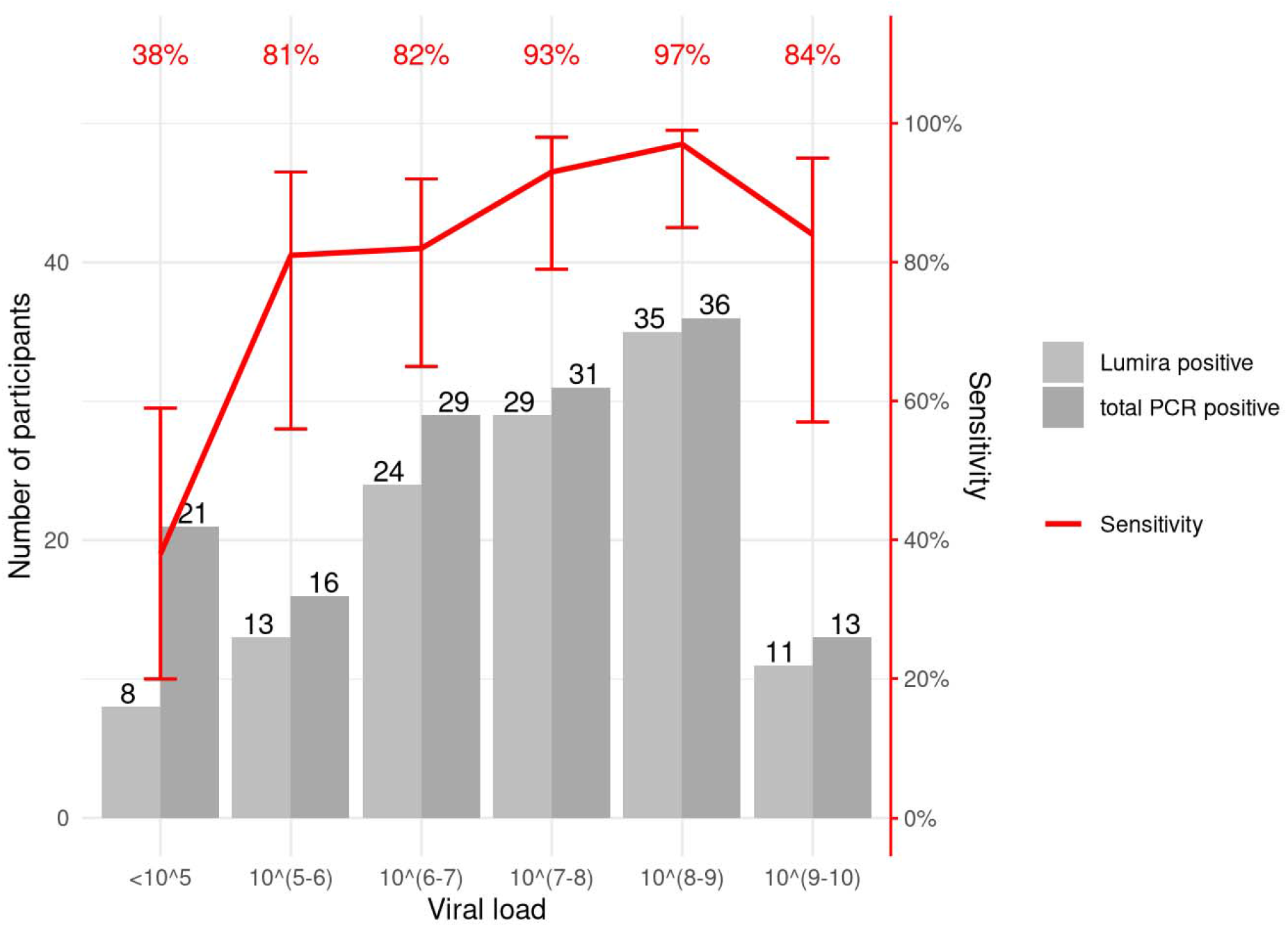
Sensitivity of LumiraDx™ compared to viral load for all RT-PCR positive participants. Viral load unit: log_10_ SARS-CoV2 RNA copies/ml

Out of 758 participants, 272 (35.9%) were asymptomatic, high-risk contacts with nine RT-PCR confirmed positive cases, resulting in a sensitivity of 77.8% with a wide confidence interval (95% CI: 45.3%-93.7%). The sensitivity among symptomatic participants was 82.5% (95% CI: 75.3%-87.9%) in comparison (Figure 2). The median viral load values of the asymptomatic cohort and the symptomatic participants were 6.3 (IQR: 3.2-7.5) and 7.4 (IQR: 3.7-9.4) log10 SARS-CoV2 RNA copies/ml, respectively. Figure 4 shows the sensitivity across asymptomatic and symptomatic participants by viral load.

**Figure 4.**
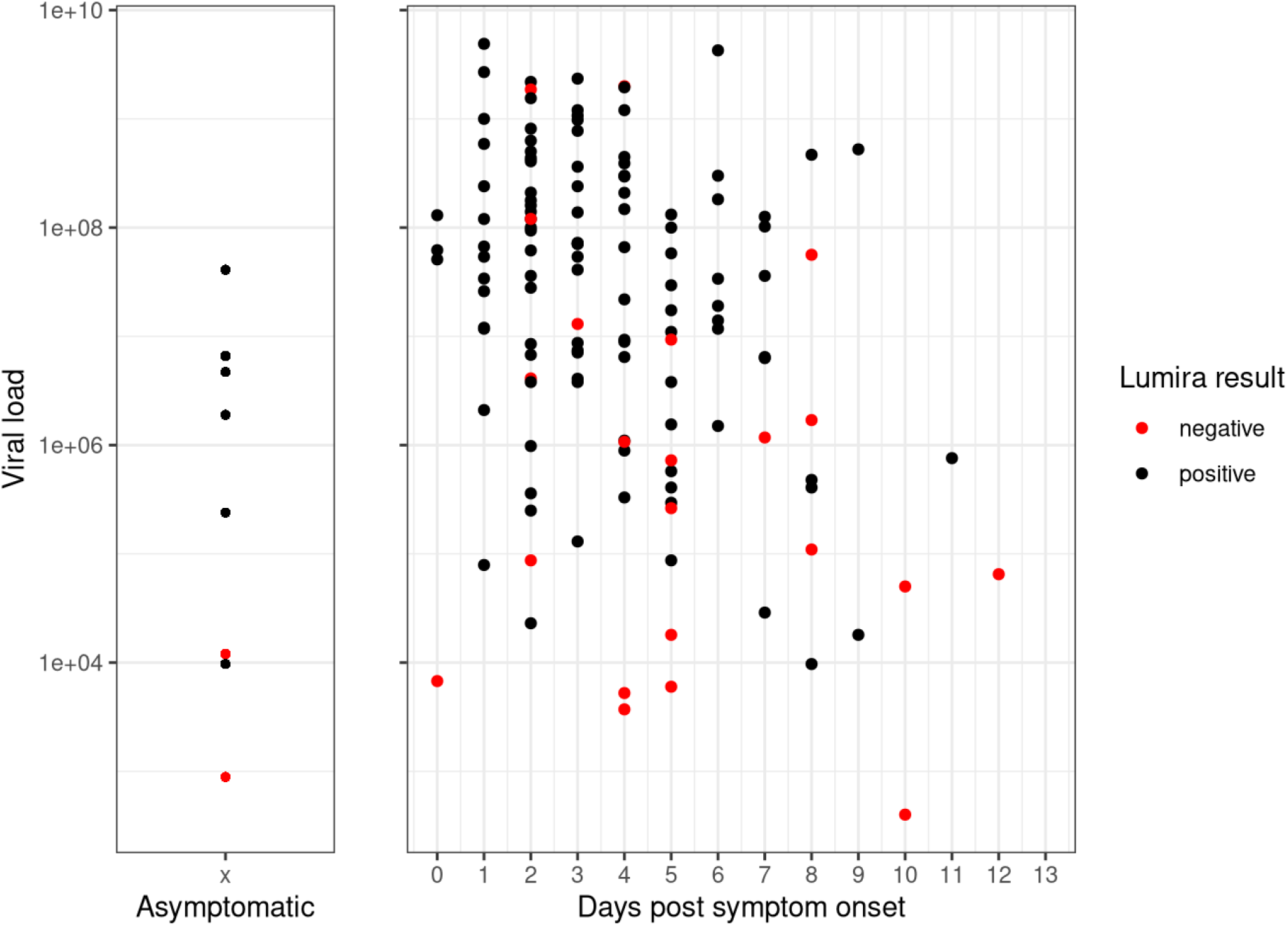
LumiraDx™ results according to viral load by presence of symptoms and days since symptom onset for all RT-PCR positive participants. Viral load unit: log_10_ SARS-CoV2 RNA copies/ml

With respect to a possible impact of duration of illness, the overall sensitivity of the Ag-RDT was found to increase slightly to 83.2% (95% CI: 75.9%-88.6%), when restricting the analysis to the manufacturer recommended 12 days post symptom onset (N=461). Sensitivity was 86.4% (95% CI: 79.1%-91.5%), with 102 of 118 RT-PCR positive cases detected, when symptom duration was 7 days or less, compared to 53.8% (95% CI: 29.1%-76.8%) for participants with a symptom duration of 8-14 days.

### Discrepant analysis

Sufficient sample volume was available to evaluate three out of three Ag-RDT positive but RT-PCR negative cases (i.e., false positive) and only nine of 26 Ag-RDT negative but RT-PCR positive cases (i.e., false negative) with an RT-PCR from the antigen buffer (thus 17 false-negative samples could not be retested). All but two out of nine false negatives tested negative. The two-testing positive on RT-PCR from the buffer showed a viral load likely below the analytical sensitivity of LumiraDx™ (4.5 and 2.6 log10 SARS-CoV2 RNA copies/ml; supplement Section (H) Table 3).

### Analytical sensitivity

To test the analytical sensitivity of LumiraDx™, a head-to-head comparison with Abbott PanBio (Chicago, Illinois, USA) and SD Biosensor Standard Q (Gyeonggi-do, Korea), using in vitro-propagated, live virus was employed. For this analysis, we tested the BavPat1 isolate produced in two different cell lines (VeroE6 and Calu-3) as well as an additional patient isolate (HD isolate). This analysis showed LumiraDx™ to have superior sensitivity to the two lateral flow assays (Figure 5) across the three tested viruses. The analytical limit of detection was estimated in the range of 2.1-55.6 PFU for LumiraDx™ which compared to 52.9 – 1,428.6 PFU and 8.8 - 238.1 PFU for the Abbott PanBio and SD Biosensor Standard Q, respectively. Of note, we observed an overall lower sensitivity in detecting *SARS-CoV-2* produced in Calu-3 cells compared to VERO cell stocks, which likely reflects higher levels of specific infectivity in virus isolated from Calu-3 cells.

**Figure 5.**
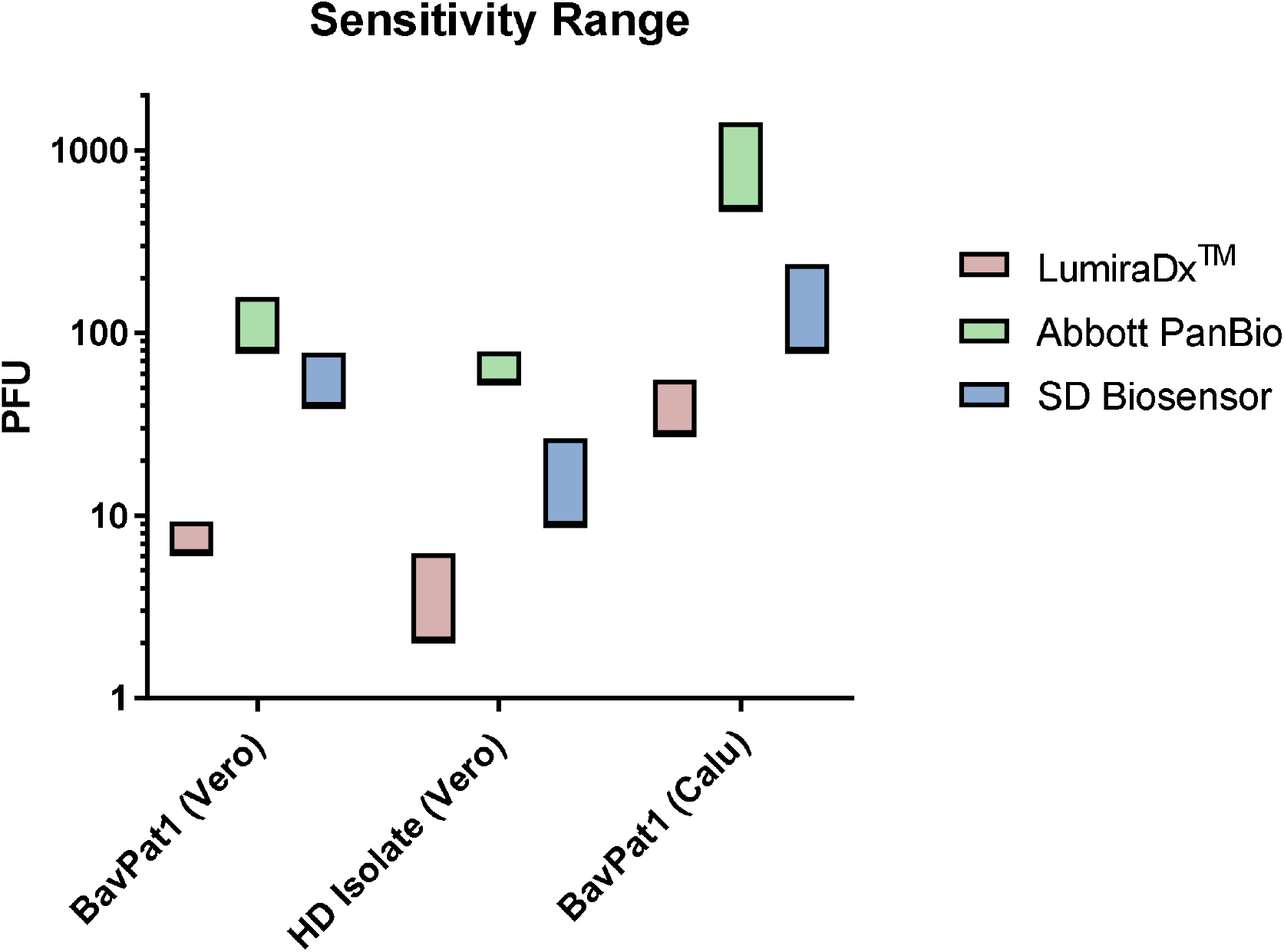
Analytical sensitivity of LumiraDx in comparison to Abbott PanBio and SD Biosensor Standard Q.

### Exclusivity testing

No cross-reactivity was detected among a wide array of respiratory pathogens and commensals tested, including the endemically circulating human coronaviruses. Detailed results of the exclusivity testing are reported in Table 4 in Section (I) in the supplementary material.

### System Usability Scale (SUS) and Ease-of-Use Assessment (EoU)

In the SUS, the LumiraDx™ scored 65 out of 100 points. The system was considered easy to use without much training required. However, the test device is not suitable for batch testing. The throughput within eight hour working shifts is a maximum of 24 tests per device, as only one sample can be processed at a time and the device requires 12 minutes analysis time, in addition to a 5-minute disinfection and drying procedure. A total storage time of five hours after sample extraction allows for testing to be organized sequentially and most likely capture the maximal throughput in a day (supplement Section (J) Figure 3 System Usability Score and Ease-of-Use assessment results).

## Discussion

This multi-centre clinical diagnostic accuracy study demonstrates a high sensitivity of 82.2% and an excellent specificity of 99.3% for the evaluated LumiraDx™ microfluidic immunofluorescence test. For participants with high viral load (≥ 7 log10 *SARS-CoV2* RNA copies/ml) who are presumably responsible for most secondary transmission, the test showed an even higher sensitivity (93.8%) (13). The assay detected most positive cases within the first week of symptoms, with a sensitivity of 86.4% (95% CI: 79.1%-91.5%). The analytical sensitivity was excellent and surpassed that of other Ag-RDTs.

The clinical sensitivity of the LumiraDx™ in this study was not better than well performing lateral flow assays that can be read with the naked eye in other studies (4, 5). The two WHO-recommended Ag-RDTs, the SD Biosensor Standard Q and Abbott PanBio demonstrated sensitivities ranging from 76.6-85.0% in our studies preceding this study (4, 14). However, as we were not able to perform the tests head-to-head to allow a direct comparison, the differences may be attributable to different phases of the pandemic with different test-positive ratios. Although the test-positive ratio during the LumiraDx™ study was higher, absolute viral load and viral load distribution were similar in our PanBio study that immediately preceded this study (4).

Another explanation for the lack of superior clinical sensitivity for LumiraDx™ could be the use of NMT sampling in contrast to NP sampling for the aforementioned lateral flow assays as NP sampling remains the gold standard (15). To what extent, however, the NMT sampling, the manufacturer recommended sample type, reduces sensitivity is unclear. A recent Swedish study suggested a reduction of 3% with NMT sampling for LumiraDx™, while a head-to head study in our hands with another Ag-RDT showed largely equivalent performance (15–17). Also, important to note is that the manufacturer recommended professional-collected NMT sampling while we asked our study participants to self-collect their sample under professional supervision. However, none of the false-negative samples in our study had observed deviations from the optimal sampling procedure, therefore, though self-collection is not recommended by the supplier and could have introduced variability, this is unlikely to have been the case. Also, recent data from other studies comparing observed self-and professional-swab sampling shows this to be unlikely as a source of large variability (16).

Nevertheless, inter-sample variability or differing distribution of virus in the nasal area during the illness course could account for some false negative results, as supported by the two samples in which virus was identified in very low amounts when antigen buffer was retested by RT-PCR, while the viral load was high in the NP sample used for RT-PCR routine testing.

Our study shows a substantially lower clinically sensitivity than the study by Drain et al. (7). While detailed comparisons are limited without access to the data of the Drain study, we would consider the participant populations to be similar, apart from the fact that we had double the percentage of asymptomatic participants. Another important difference of note is in the swabs used. While the study by Drain et al. utilized a flocked swab, we utilized a standard rayon swab, supplied by the manufacturer, which might contribute to less virus being captured in our study (18).

The analytical results of the study confirm the claim of the manufacturer to have superior sensitivity over the two other Ag-RDTs studied. Importantly, values obtained for the PanBio and Standard Q in our study were similar to previous studies (19). Reasons for the high analytical sensitivity not translating into high clinical sensitivity could relate to problems in the extraction of the virus. However, the samples with high viral load that went undetected did not have any particular characteristics (consistency, blood contamination), leaving the reasons for the possible extraction problems unexplained. Another consideration could be immune escape due to viral variants not recognized by the antibodies included in the test. This could explain variable performance between two geographically distinct sites (in the Pacific North West of the USA in the study by Drain et al. (7) and Germany). However, this appears very unlikely given the multiple antibodies used in the test that target the nucleocapsid, which is a rather conserved target even in recent strain variants (20). While a hook or prozone effect could be considered, it appears unlikely in light of the samples type and has not been shown in prior experiments by the manufacturer (https://www.lumiradx.com/uk-en/what-we-do/diagnostics/test-technology/antigen-test). Thus, further studies are needed to explain the lower clinical sensitivity of LumiraDx™ for some high viral load samples and the differences observed in clinical and analytical sensitivity.

Nevertheless, it is to note, that the RT-PCR reference standard also has its limitations, as it is not always a meaningful test when considering viable virus and risk of transmission (21). This is also demonstrated by a recent study of an Ag-RDT in the USA, where the sensitivity was 92.6% in symptomatic individuals when compared to viral culture versus 64.2% when compared to RT-PCR (22). Thus, using the RT-PCR reference standard, we might have underestimated the performance of the Ag-RDT when it comes to detection of viable virus.

We also acknowledge that the Lumira assay is only recommended for symptomatic patients. Sensitivity was slightly higher among symptomatic patients (82.5%, 95% CI:75.3%-87.9%) as compared to asymptomatic individuals (77.8%, 45.3%-93.7%), with overlapping confidence intervals. This was to be expected from the lower viral load observed in the limited number of asymptomatic positive cases. As cohort studies have shown the viral load kinetics between asymptomatic and symptomatic infections to be similar, the findings of the lower viral load and thus lower sensitivity observed in our study is most likely attributable to chance or to capturing asymptomatic patients too early (maybe also prior to symptom development). Further need for studies remain to better understand the performance in asymptomatic patients (23).

The excellent clinical specificity of the LumiraDx™ was confirmed by the analytical exclusivity testing. Our findings indicate no cross-reactivity with the eleven tested pathogens or commensals that would lead to false-positive Ag-RDT results. Assuming a *SARS-CoV-2* prevalence of 1%, a test with the performance of the LumiraDx™ would result in a positive predictive value (PPV) of 45.8% and a negative predictive value (NPV) of 99.8%. At a prevalence of 3%, PPV would be 78.7% and NPV 99.5%.

The NMT sampling was considered highly favourable based on participants feedback in comparison to the NP sampling. In addition, NMT self-sampling under professional supervision would enable a higher sampling throughput, requiring fewer medical personnel and less protective equipment used (e.g., the sampling could be observed through a window). Another very favourable aspect of the test is the possibility for results to be reported automatically and recorded digitally to be uploaded in information systems, which is an important characteristic when it comes to surveillance and supply chain management. On the other hand, the device-based assay does not allow for batch testing and has a limited throughput. Thus, our observations show the LumiraDx™ to likely be most useful in an environment where persons to be tested arrive over time, such as a medical office or emergency room. The device-based LumiraDx™ system may also lend itself to simultaneous testing for several respiratory pathogens from the same patient sample, which could facilitate the workup of individuals presenting with non-specific respiratory symptoms, e.g., during the influenza season.

Overall, our study has several strengths. The study sites in Heidelberg and Berlin enrolled participants representative of the current pandemic observed in Germany. The study participants represented the age groups above 18 most commonly presenting for testing in an ambulatory setting and including asymptomatic and symptomatic participants across the two study sites, making the findings generalizable to ambulatory test settings across Germany. The accuracy of the Ag-RDT was evaluated using several measures, including clinical accuracy, analytical sensitivity, and exclusivity. In addition, the ease-of-use assessment especially designed for this study, in combination with the standardized SUS, emphasized important points regarding the operationalization of the test. Finally, this study was conducted in point-of-care settings, representing the general diagnostic challenges of near patient testing.

However, our study also has limitations. The population was preselected by the local health department’s testing criteria based on national guidelines. As pandemic dynamics change, these criteria are regularly updated, yet no significant differences were observed in enrolled study population groups over the study time in terms of comparability. The sub-analyses by Ct-value have limited comparability as three different RT-PCR standards were used for this study, using different assay targets, reagents, and technical equipment. To account for possible variability across tests in Ct-values, viral load was calculated using a standardized calibrated method and used for the comparisons.

In conclusion, we demonstrate that the LumiraDx™ assay has comparable clinical sensitivity and higher analytical sensitivity than WHO-approved lateral-flow Ag-RDTs, paired with excellent specificity and operational characteristics suitable for low-throughput settings.

## Data Availability

We are currently working on the availability of the dataset in a file repository and will make a link available as soon as we have completed the process

## Acknowledgments

Angelika Sandritter, Alexander Syring, and the KTS company performing the routine testing at the Heidelberg site

## Study Team

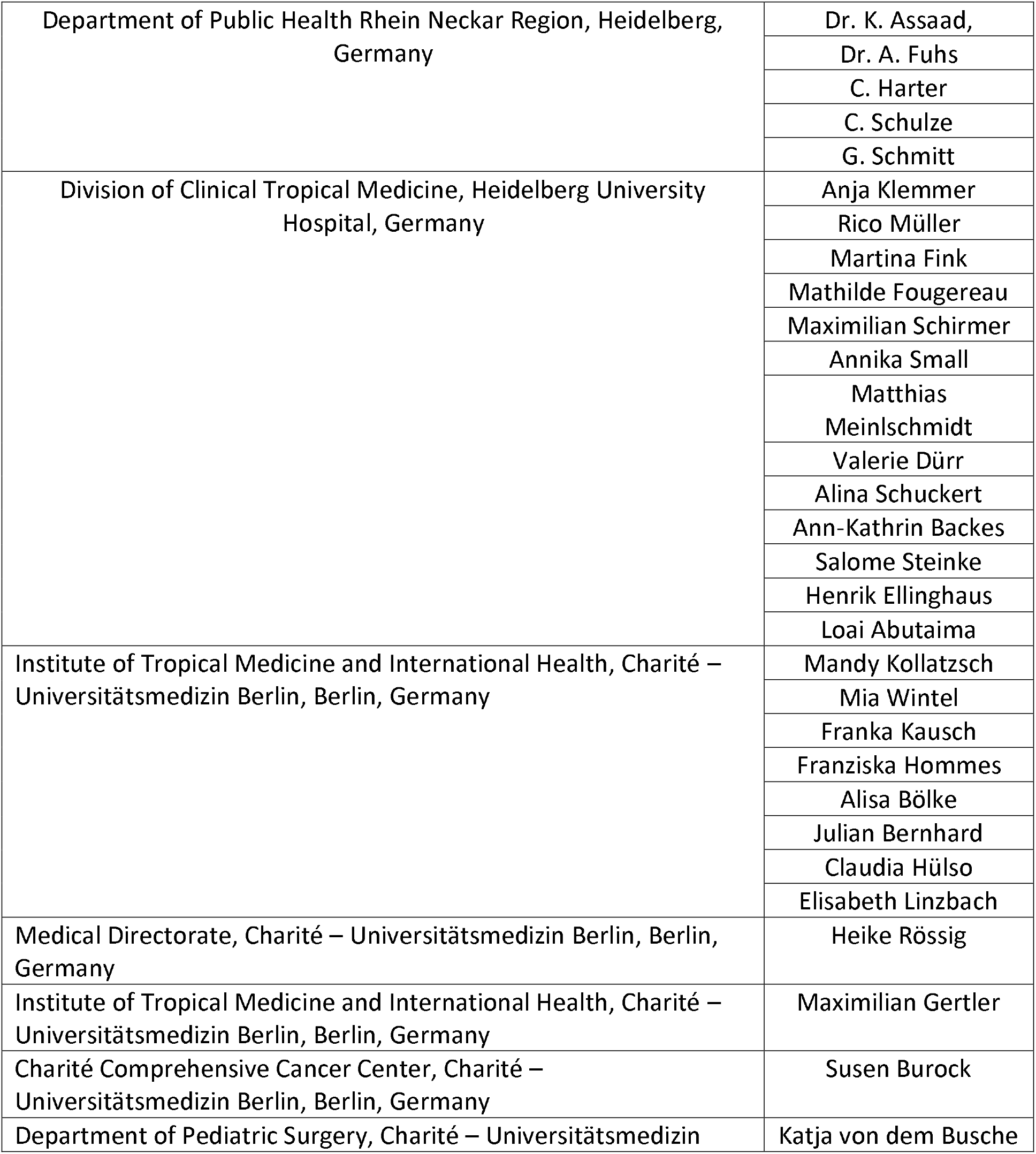
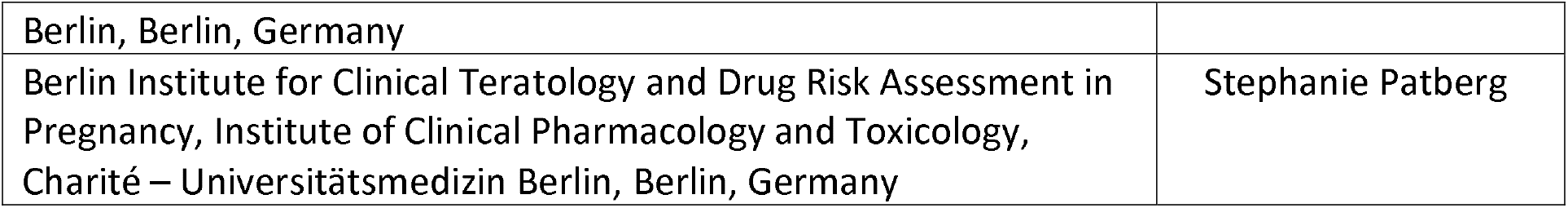

## References

1. Corman VM, Landt O, Kaiser M, Molenkamp R, Meijer A, Chu DK, Bleicker T, Brunink S, Schneider J, Schmidt ML, Mulders DG, Haagmans BL, van der Veer B, van den Brink S, Wijsman L, Goderski G, Romette JL, Ellis J, Zambon M, Peiris M, Goossens H, Reusken C, Koopmans MP, Drosten C. 2020. Detection of 2019 novel coronavirus (2019-nCoV) by real-time RT-PCR. Euro Surveill 25.

2. Holt E. 2020. Slovakia to test all adults for SARS-CoV-2. The Lancet 396:1386–1387.

3. Berger A, Ngo Nsoga M-T, Perez Rodriguez FJ, Abi Aad Y, Sattonnet P, Gayet-Ageron A, Jaksic C, Torriani G, Boehm E, Kronig I, Sacks JA, de Vos M, Jacquerioz-Bausch F, Chappuis F, Kaiser L, Schibler M, Eckerle I. 2020. Diagnostic accuracy of two commercial SARS-CoV-2 Antigen-detecting rapid tests at the point of care in community-based testing centers. medRxiv doi:10.1101/2020.11.20.20235341:2020.11.20.20235341.

4. Krüger LJ, Gaeddert M, Tobian F, Lainati F, Gottschalk C, Klein JAF, Schnitzler P, Kräusslich HG, Nikolai O, Lindner AK, Mockenhaupt FP, Seybold J, Corman VM, Drosten C, Pollock NR, Knorr B, Welker A, de Vos M, Sacks JA, Denkinger CM. 2020. Evaluation of the accuracy and ease-of-use of Abbott PanBio - A WHO emergency use listed, rapid, antigen-detecting point-of-care diagnostic test for SARS-CoV-2. medRxiv doi:10.1101/2020.11.27.20239699:2020.11.27.20239699.

5. Krüger LJ, Gaeddert M, Köppel L, Brümmer LE, Gottschalk C, Miranda IB, Schnitzler P, Kräusslich HG, Lindner AK, Nikolai O, Mockenhaupt FP, Seybold J, Corman VM, Drosten C, Pollock NR, Cubas-Atienzar AI, Kontogianni K, Collins A, Wright AH, Knorr B, Welker A, de Vos M, Sacks JA, Adams ER, Denkinger CM. 2020. Evaluation of the accuracy, ease of use and limit of detection of novel, rapid, antigen-detecting point-of-care diagnostics for SARS-CoV-2. medRxiv doi:10.1101/2020.10.01.20203836:2020.10.01.20203836.

6. Basiri A, Heidari A, Nadi MF, Fallahy MTP, Nezamabadi SS, Sedighi M, Saghazadeh A, Rezaei N. 2020. Microfluidic devices for detection of RNA viruses. Reviews in medical virology doi:10.1002/rmv.2154:e2154–e2154.

7. Drain PK, Ampajwala M, Chappel C, Gvozden AB, Hoppers M, Wang M, Rosen R, Young S, Zissman E, Montano M. 2020. A rapid, high-sensitivity SARS-CoV-2 nucleocapsid immunoassay to aid diagnosis of acute COVID-19 at the point of care. medRxiv doi:10.1101/2020.12.11.20238410:2020.12.11.20238410.

8. Cohen JF, Korevaar DA, Altman DG, Bruns DE, Gatsonis CA, Hooft L, Irwig L, Levine D, Reitsma JB, de Vet HCW, Bossuyt PMM. 2016. STARD 2015 guidelines for reporting diagnostic accuracy studies: explanation and elaboration. BMJ Open 6:e012799.

9. Harris PA, Taylor R, Thielke R, Payne J, Gonzalez N, Conde JG. 2009. Research electronic data capture (REDCap)--a metadata-driven methodology and workflow process for providing translational research informatics support. J Biomed Inform 42:377–81.

10. Klein S, Cortese M, Winter SL, Wachsmuth-Melm M, Neufeldt CJ, Cerikan B, Stanifer ML, Boulant S, Bartenschlager R, Chlanda P. 2020. SARS-CoV-2 structure and replication characterized by in situ cryo-electron tomography. Nature Communications 11:5885.

11. Bangor A, Kortum PT, Miller JT. 2008. An Empirical Evaluation of the System Usability Scale. International Journal of Human–Computer Interaction 24:574–594.

12. Altman DG, Bland JM. 1994. Diagnostic tests. 1: Sensitivity and specificity. BMJ 308:1552.

13. Meyerowitz EA, Richterman A, Gandhi RT, Sax PE. 2020. Transmission of SARS-CoV-2: A Review of Viral, Host, and Environmental Factors. Annals of internal medicine doi:10.7326/M20-5008:M20-5008.

14. Lindner AK, Nikolai O, Rohardt C, Kausch F, Wintel M, Gertler M, Burock S, Hörig M, Bernhard J, Tobian F, Gaeddert M, Lainati F, Corman VM, Jones TC, Sacks JA, Seybold J, Denkinger CM, Mockenhaupt FP. 2021. SARS-CoV-2 patient self-testing with an antigen-detecting rapid test: a head-to-head comparison with professional testing. medRxiv doi:10.1101/2021.01.06.20249009:2021.01.06.20249009.

15. Scandinavian evaluation of laboratory equipment for point of care testing. 2020. LumiraDx: A system for detection of SARS-CoV-2 antigen and antibodies, PT (INR) and D-Dimer manufactured by LumiraDx UK Ltd - An evaluation of the detection of SARS-CoV-2 antigen.

16. Lindner AK, Nikolai O, Kausch F, Wintel M, Hommes F, Gertler M, Kruger LJ, Gaeddert M, Tobian F, Lainati F, Koppel L, Seybold J, Corman VM, Drosten C, Hofmann J, Sacks JA, Mockenhaupt FP, Denkinger CM. 2020. Head-to-head comparison of SARS-CoV-2 antigen-detecting rapid test with self-collected anterior nasal swab versus professional-collected nasopharyngeal swab. Eur Respir J doi:10.1183/13993003.03961-2020.

17. Lindner AK, Nikolai O, Rohardt C, Burock S, Hülso C, Bölke A, Gertler M, Krüger LJ, Gaeddert M, Tobian F, Lainati F, Seybold J, Jones TC, Hofmann J, Sacks JA, Mockenhaupt FP, Denkinger CM. 2021. Head-to-head comparison of SARS-CoV-2 antigen-detecting rapid test with professional-collected nasal versus nasopharyngeal swab. European Respiratory Journal doi:10.1183/13993003.04430-2020:2004430.

18. Lee RA, Herigon JC, Benedetti A, Pollock NR, Denkinger CM. 2020. Performance of Saliva, Oropharyngeal Swabs, and Nasal Swabs for SARS-CoV-2 Molecular Detection: A Systematic Review and Meta-analysis. medRxiv doi:10.1101/2020.11.12.20230748:2020.11.12.20230748.

19. Corman VM, Haage VC, Bleicker T, Schmidt ML, Mühlemann B, Zuchowski M, Jó Lei WK, Tscheak P, Möncke-Buchner E, Müller MA, Krumbholz A, Drexler JF, Drosten C. 2020. Comparison of seven commercial SARS-CoV-2 rapid Point-of-Care Antigen tests. medRxiv doi:10.1101/2020.11.12.20230292:2020.11.12.20230292.

20. Haynes WA, Kamath K, Lucas C, Shon J, Iwasaki A. 2021. Impact of B.1.1.7 variant mutations on antibody recognition of linear SARS-CoV-2 epitopes. medRxiv doi:10.1101/2021.01.06.20248960:2021.01.06.20248960.

21. Sun J, Xiao J, Sun R, Tang X, Liang C, Lin H, Zeng L, Hu J, Yuan R, Zhou P, Peng J, Xiong Q, Cui F, Liu Z, Lu J, Tian J, Ma W, Ke C. 2020. Prolonged Persistence of SARS-CoV-2 RNA in Body Fluids. Emerging Infectious Disease journal 26:1834.

22. Prince-Guerra J AO, Nolen L, et al. 2020. Evaluation of Abbott BinaxNOW Rapid Antigen Test for SARS-CoV-2 Infection at Two Community-Based Testing Sites — Pima County, Arizona. MMWR Morb Mortal Wkly Rep 2021 70:100–105.

23. Kissler SM, Fauver JR, Mack C, Olesen SW, Tai C, Shiue KY, Kalinich CC, Jednak S, Ott IM, Vogels CBF, Wohlgemuth J, Weisberger J, DiFiori J, Anderson DJ, Mancell J, Ho DD, Grubaugh ND, Grad YH. 2020. SARS-CoV-2 viral dynamics in acute infections. medRxiv doi:10.1101/2020.10.21.20217042:2020.10.21.20217042.

