## Supplementary Material for "Evaluation of accuracy, exclusivity, limit-of-detection and ease-of-use of LumiraDx™-Antigen-detecting point-of-care device for *SARS-CoV-2*"

#### (A) Section: Questionnaire for study participants

**We invite you to participate in this survey. The survey serves to understand the diagnostic process and the disease and factors related to SARS-CoV-2 (novel coronavirus) infection.**

**Your answers will be kept strictly confidential and will not have a negative impact on your care. Participation in the study is voluntary and you have the option to skip questions that you do not want to answer.**

**The survey is expected to take 15-20 minutes. Thank you for your understanding and cooperation!**

| | |
|---|---|
| Postal code | *(Free text)* |
| Gender | ○ Male<br>○ Female<br>○ Diverse |
| How tall are you (in centimetres)? | *(Free text)* |
| How much do you weight (in kilograms)? | *(Free text)* |

**Symptoms that you attribute to the possible COVID-19**

| | |
|---|---|
| Did you have any symptoms of possible COVID-19 on the day of the test? | ○ No<br>○ Yes |
| Increased temperature / fever? | ○ No<br>○ Yes |
| Did you measure your fever? | ○ No<br>○ Yes |
| Highest temperature (in Celsius) | *(Free text)* |
| Cough | ○ No<br>○ Yes |
| Do you have a productive cough? | ○ No<br>○ Yes |
| Sore throat | ○ No<br>○ Yes |
| Shortness of breath | ○ No<br>○ Yes |
| Muscle pain / Body aches | ○ No<br>○ Yes |

| | |
|---|---|
| Fatigue | ○ No<br>○ Yes |
| Headache | ○ No<br>○ Yes |
| Runny nose | ○ No<br>○ Yes |
| Chest pain | ○ No<br>○ Yes |
| Diarrhea | ○ No<br>○ Yes |
| Nausea / vomiting | ○ No<br>○ Yes |
| Loss of taste or smell | ○ No<br>○ Yes |
| Other | ○ No<br>○ Yes |
| If yes, please specify | *(Free text)* |
| The earliest onset of symptoms attributed to possible COVID-19 | *(Day / Month / Year)* |
| How sick did you feel on the day of the test? | ○ Normal unrestricted activity as before the illness<br>○ Restriction with physical exertion, but able to walk; light physical work or work while sitting, e.g., light housework or office work, possible<br>○ Able to walk, self-sufficiency possible, but not able to work; can get up more than 50% of the waking time<br>○ Only limited self-sufficiency possible; 50% or more of the waking time tied to bed or chair<br>○ Completely in the need of care, no self-sufficiency possible; completely tied to bed or chair |
| Did you previously test negative within the last 10 days? | ○ No<br>○ Yes |
| If yes, when | *(Day / Month / Year)* |
| If yes, where | ○ University clinic – inpatient<br>○ University clinic – outpatient<br>○ Drive-in |

| | |
|---|---|
| | ○ Other |
| Other, please specify | *(Free text)* |
| Do you know where you might have been infected with COVID-19? | ○ Household contact<br>○ Social contact<br>○ Work contact<br>○ Contact in university / school / kindergarten from you or a child in the family<br>○ Travel to risk area<br>○ Do not know<br>○ Other |
| Risk area | *(Free text)* |
| Other: please describe | *(Free text)* |

**Do you have any pre-existing conditions?**

| | |
|---|---|
| Which of the following lung disease(s) do you have? | ○ Asthma<br>○ Chronic Obstructive Pulmonary Disease – COPD<br>○ Breathing disorders during sleep<br>○ Obstructive Sleep Apnea – OSAS<br>○ Interstitial Lung Disease<br>○ Lung Cancer<br>○ Other<br>○ None |
| What other lung diseases do you have? | *(Free text)* |
| Cardiovascular diseases (e.g. hypertension, stroke, etc.) | ○ No<br>○ Yes |
| Chronic kidney disease | ○ No<br>○ Yes |
| Diabetes | ○ No<br>○ Yes |
| Autoimmune Disease (e.g. Rheumatoid Arthritis, MS) | ○ No<br>○ Yes |
| HIV | ○ No<br>○ Yes |
| Overweight | ○ No<br>○ Yes |
| Other, please specify | *(Free text)* |

### (B) Section: System Usability Scale (SUS)

"Evaluation of the performance of novel rapid diagnostics for SARS-CoV-2 at point-of-care"

***System Usability Scale (SUS)***

© Digital Equipment Corporation, 1986 adapted format, version 1.0: 02/05/20

Name of the test: ________________________________________________________________

User identifier and study site: _________________________ Date: _______________________

| | Strongly disagree 1 | 2 | 3 | 4 | Strongly agree 5 |
|---|---|---|---|---|---|
| 1. I think that I would like to use this system frequently | ☐ | ☐ | ☐ | ☐ | ☐ |
| 2. I found the system unnecessarily complex | ☐ | ☐ | ☐ | ☐ | ☐ |
| 3. I thought the system was easy to use | ☐ | ☐ | ☐ | ☐ | ☐ |
| 4. I think that I would need the support of a technical person to be able to use this system | ☐ | ☐ | ☐ | ☐ | ☐ |
| 5. I found the various functions in this system were well integrated | ☐ | ☐ | ☐ | ☐ | ☐ |
| 6. I thought there was too much inconsistency in this system | ☐ | ☐ | ☐ | ☐ | ☐ |
| 7. I would imagine that most people would learn to use this system very quickly | ☐ | ☐ | ☐ | ☐ | ☐ |
| 8. I found the system very cumbersome to use | ☐ | ☐ | ☐ | ☐ | ☐ |
| 9. I felt very confident using the system | ☐ | ☐ | ☐ | ☐ | ☐ |
| 10. I needed to learn a lot of things before I could get going with this system | ☐ | ☐ | ☐ | ☐ | ☐ |

***Using SUS***

The SU scale is generally used after the respondent has had an opportunity to use the system being evaluated, but before any debriefing or discussion takes place. Respondents should be asked to record their immediate response to each item, rather than thinking about items for a long time.

All items should be checked. If a respondent feels that they cannot respond to a particular item, they should mark the center point of the scale.

#### (C) Section: Ease-of-Use Assessment (EoU)

"Evaluation of the performance of novel rapid diagnostics for SARS-CoV-2 at point-of-care"

Thank you for your time to answer this questionnaire (about 20 minutes).

*Your input is very valuable!*

***OVERALL QUESTIONS***

**1. User identifier**

**2. Date of filling the questionnaire**

**3. In which country do you currently work?**

**4. At which facility / study site do you currently work?**

*Mark only one oval.*

- ⬭ Reilingen (Heidelberg)
- ⬭ Berlin
- ⬭ Liverpool
- ⬭ Macae CTC
- ⬭ Marica (Guapi)
- ⬭ UFRJ

**5. Which test are you assessing?**

*Mark only one oval.*

- ⬭ Coris Bioconcept Respi Strip
- ⬭ Bioeasy FIA
- ⬭ Bioeasy Colloidal Gold
- ⬭ SD Biosensor Standard F (Flourescence)
- ⬭ SD Biosensor Standard Q
- ⬭ Rapigen Biocredit Colloidal Gold

- [ ] Abbott Panbio
- [ ] LumiraDx

**6. Approximately how many times did you perform this test?**

*Mark only one oval.*

- [ ] Only observed use
- [ ] < 10
- [ ] 10 – 100
- [ ] > 100

**7. Approximately how many times did you observe the use of this test (not performed yourself)?**

*Mark only one oval.*

- [ ] < 10
- [ ] 10 – 50
- [ ] 50 – 100
- [ ] > 100

**8. What is your profession?**

______________________________________________________________________

**9. How many years of laboratory experience do you have?**

______________________________________________________________________

**10. How many years of working experience in limited resource settings do you have?**

______________________________________________________________________

11. **How much experience do you have with interpreting the results of lateral flow tests or rapid diagnostics (e.g. for HIV, malaria, pregnancy)?**

Please note that we refer here to your experience with INTERPRETING the test results. If you do not conduct the test yourself, but do inform patients about the test results, we also consider that as experience with INTERPRETING the test results.

*Mark only one oval.*

- None
- < 1 year
- 1 – 3 years
- > 3 years

***TEST SPECIFIC QUESTIONS***

*TRAINING*

12. **How satisfied were you with the following components of the test training?**

*Mark only one oval per row.*

| | Very satisfied | Satisfied | Neither | Dissatisfied | Very dissatisfied |
|---|---|---|---|---|---|
| Instructions for Use | | | | | |
| Standard Operating Procedures | | | | | |
| Face to face demonstration | | | | | |

13. **What additional materials (if any) do you think should be provided as part of the training?**

- None
- Other: ______________________________________________

14. **How long should be the training of this test?**

*Mark only one oval.*

- Self-explanatory, no need for training
- 1 - 2 hours

- [ ] 2 - 4 hours
- [ ] Half a day
- [ ] Full day

**15. Do you consider proficiency testing necessary?**

Proficiency testing as in assessing the user's performance or ability to run the test following the training.

*Mark only one oval.*

- [ ] Yes
- [ ] No
- [ ] Other: : ____________________________________________

**16. Please comment here on the need for proficiency testing**

____________________________________________

**17. Do you think that a company training is necessary?**

- [ ] Yes
- [ ] No
- [ ] Other: : ____________________________________________

**18. Please comment on having the company come to do an in-person training.**

____________________________________________

**19. After how many of these tests do you feel you could perform the test on your own (having access to the training material)?**

*Mark only one oval.*

- [ ] 1 – 2 tests
- [ ] 3 – 5 tests

( ) 6 – 10 tests

( ) > 10 tests

*<u>ASSESSMENT OF TEST COMPONENTS</u>*

**20. How satisfied are you with the quality of each of the components in the Test Strip Carton (in terms of ease of use and fit for purpose)?**

*Mark only one oval per row*

| | Very satisfied | Satisfied | Neither | Dissatisfied | Very dissatisfied |
|---|---|---|---|---|---|
| External paper box of kit | ( ) | ( ) | ( ) | ( ) | ( ) |
| Extraction tube | ( ) | ( ) | ( ) | ( ) | ( ) |
| Filter cap | ( ) | ( ) | ( ) | ( ) | ( ) |
| Test cartridge / device | ( ) | ( ) | ( ) | ( ) | ( ) |
| Test cartridge packing / pouch | ( ) | ( ) | ( ) | ( ) | ( ) |

**21. Which kit component(s) should be improved in your opinion (if any)?**
*Please specify why and how*

________________________________________________________________________________

**22. Overall, how satisfied are you with the kit components?**

*Mark only one oval.*

| | 1 | 2 | 3 | 4 | 5 | |
|---|---|---|---|---|---|---|
| Very satisfied | ( ) | ( ) | ( ) | ( ) | ( ) | Very dissatisfied |

**23. How satisfied are you with the overall design of the test strip in terms of the following features?**

*Mark only one oval per row.*

| | Very satisfied | Satisfied | Neither | Dissatisfied | Very dissatisfied |
|---|---|---|---|---|---|
| Size of test strip | ( ) | ( ) | ( ) | ( ) | ( ) |
| Size of the well to add sample mix | ( ) | ( ) | ( ) | ( ) | ( ) |

**24. How satisfied are you with the quality of the quality controls in terms of ease of use and fit for purpose?**

1 2 3 4 5

| | | | | | | |
|---|---|---|---|---|---|---|
| Very satisfied | ⬭ | ⬭ | ⬭ | ⬭ | ⬭ | Very dissatisfied |

**25. How useful do you find the inclusion of a positive quality control?**

| | 1 | 2 | 3 | 4 | 5 | |
|---|---|---|---|---|---|---|
| Very useful | ⬭ | ⬭ | ⬭ | ⬭ | ⬭ | Not useful |

**26. How useful do you find the inclusion of a negative quality control?**

| | 1 | 2 | 3 | 4 | 5 | |
|---|---|---|---|---|---|---|
| Very useful | ⬭ | ⬭ | ⬭ | ⬭ | ⬭ | Not useful |

**27. Please comment on the added value and quality of the quality controls.**

___________________________________________________________________________

**28. How satisfied are you with the quality of the reader in terms of its ease of use and fit for purpose?**

| | 1 | 2 | 3 | 4 | 5 | |
|---|---|---|---|---|---|---|
| Very satisfied | ⬭ | ⬭ | ⬭ | ⬭ | ⬭ | Very dissatisfied |

***29.*** **Overall, how satisfied are you with the reader?**

| | 1 | 2 | 3 | 4 | 5 | |
|---|---|---|---|---|---|---|
| Very satisfied | ⬭ | ⬭ | ⬭ | ⬭ | ⬭ | Very dissatisfied |

**30. Which component(s) of the reader (if applicable) should be improved in your opinion (if any)? Please specify why and how.**

___________________________________________________________________________

*ASSESSMENT OF TEST*

**31. Please determine the difficulty of the following steps:**
Please consider your day-to-day/routine workload (or that of the people in the lab/area where this test could be implemented) to answer this question. Please leave any steps blank if not applicable.

*Mark only one oval per row.*

| | Very easy | Easy | Neither | Difficult | Very difficult |
|---|---|---|---|---|---|
| a) Check expiry date | ⬭ | ⬭ | ⬭ | ⬭ | ⬭ |

| | | | | | |
|---|---|---|---|---|---|
| b) Label the extraction tube with patient identifier | ⬭ | ⬭ | ⬭ | ⬭ | ⬭ |
| c) Open the extraction tube by removing the seal | ⬭ | ⬭ | ⬭ | ⬭ | ⬭ |
| d) Insert the swab into the tube | ⬭ | ⬭ | ⬭ | ⬭ | ⬭ |
| e) Ease of swab extraction procedure | ⬭ | ⬭ | ⬭ | ⬭ | ⬭ |
| f) Ability to perform swab extraction procedure consistently | ⬭ | ⬭ | ⬭ | ⬭ | ⬭ |
| g) Ability to maintain cleanliness of ancillary devices (e.g. pipette) in order to avoid cross contamination | ⬭ | ⬭ | ⬭ | ⬭ | ⬭ |
| h) Remove the test cartridge from the pouch | ⬭ | ⬭ | ⬭ | ⬭ | ⬭ |
| i) Ease of transferring sample onto device | ⬭ | ⬭ | ⬭ | ⬭ | ⬭ |
| j) Ease of transferring exact quantity into the sample well | ⬭ | ⬭ | ⬭ | ⬭ | ⬭ |
| k) Ease of using instrument | ⬭ | ⬭ | ⬭ | ⬭ | ⬭ |
| l) Trouble shooting | ⬭ | ⬭ | ⬭ | ⬭ | ⬭ |

**32. How satisfied are you with the logical sequence of steps?**

*Mark only one oval.*

| | 1 | 2 | 3 | 4 | 5 | |
|---|---|---|---|---|---|---|
| Very satisfied | ⬭ | ⬭ | ⬭ | ⬭ | ⬭ | Very dissatisfied |

**33. Overall, how difficult did you find the steps?**

*Mark only one oval.*

| | 1 | 2 | 3 | 4 | 5 | |
|---|---|---|---|---|---|---|
| Very easy | ⬭ | ⬭ | ⬭ | ⬭ | ⬭ | Very difficult |

**34.** **In general, how satisfied are you with the time relevant components (ex. are the steps relatively short, are there many specific timed steps that make it hard to keep track)?**

*Mark only one oval.*

| | 1 | 2 | 3 | 4 | 5 | |
|---|---|---|---|---|---|---|
| Very satisfied | ⬭ | ⬭ | ⬭ | ⬭ | ⬭ | Very dissatisfied |

**35. Please assess the time relevant components of the test.**

*Mark only one oval per row.*

| | ≤ 2 min | 3 to 5 min | 6 to 10 min | > 10 min |
|---|---|---|---|---|
| Pre analytic time (i.e. from when you get the swab to when you start incubation) | ⬭ | ⬭ | ⬭ | ⬭ |
| Incubation time (how long do you have to wait to get the results once you add your sample to the strip) | ⬭ | ⬭ | ⬭ | ⬭ |
| Analytic time (time needed to analyse the results) | ⬭ | ⬭ | ⬭ | ⬭ |

**36.** **In your opinion, about how many patients could be tested with this test in an 8-hour day (with one instrument and one sample method only)?**

*Mark only one oval per row.*

- ⬭ < 10
- ⬭ 10 – 50
- ⬭ 50 – 100
- ⬭ > 100

**37.** **Please comment here if you see any potential issues or room for improvement.**

______________________________________________________________________________

**38.** **How did you find the results read-out in the followingareas?**

*Mark only one oval per row.*

| | Very easy | Easy | Neither | Difficult | Very difficult |
|---|---|---|---|---|---|
| a) Read-out from Reader | ⬭ | ⬭ | ⬭ | ⬭ | ⬭ |
| b) Interpretation of the test result | ⬭ | ⬭ | ⬭ | ⬭ | ⬭ |

**39. Do you foresee any issues with reading these results considering the lighting conditions in the settings you currently work or have experience with?**

*Mark only one oval.*

- ⬭ Yes (please explain below)
- ⬭ No

**If yes, please explain here**

_______________________________________________________________________________

*OVERALL ASSESMMENT*

**40. Overall, how did you find the use of this rapid COVID-19 diagnostic tool:**

*Mark only one oval.*

| | 1 | 2 | 3 | 4 | 5 | |
|---|---|---|---|---|---|---|
| Very easy | ⬭ | ⬭ | ⬭ | ⬭ | ⬭ | Very difficult |

**41. Please comment here on the use:**

_______________________________________________________________________________

**42. Which option(s) do you consider feasible in your setting?**

- ⬭ Sequential testing (run tests one by one) ONLY
- ⬭ Sequential testing AND batch testing (run multiple tests at the same time)

***43.*** **Which aspect(s) of this test could cause difficulties in its day-to-day use?** *Tick all that apply.*

- Hands-on time
- Tatal assay time to result
- Batch processing
- Throughput
- Test results interpretation
- Overall number of steps
- Time sensitive steps
- Cartridge design
- Quality of material
- Training requirements
- Storage conditions and stability
- Waste management requirements
- I don’t know
- None, I see no barriers for implementation

**44. Please give a short explanation for each of the aspects you selected above e.g. what could be the challenges in the day-to-day use:**

______________________________________________________________________________

*SETTINGS UF USE*

**45. Do you see this test being used in its current form in your setting in your country?**

*Mark only one oval.*

- Yes (please explain below)
- No (please explain below)
- I don’t know

Please elaborate:

___

**46. If yes, at which health care level(s) do you see this test being implemented in your country**

*Tick all that apply.*

- Family doctor / General physician
- Peripheral hospital / lab
- Reference hospital / lab
- At a testing site operated by traines staff without specific laboratory expertise

**47. If you don't see this test being used in its current form, which aspects should be changed to make it suitable for use in your setting in your country:**

___

**48. If you don't see this test being used in its current form, which aspects should be changed to make it suitable for use in your setting in low- and middle-income countries?**

___

**49. Anything else you would like to add?**

___

#### THANK YOU VERY MUCH

___

### (D) Figure 1: Matrix for Ease-of-Use Assessment

| | Timestamp | green | green | green | yellow | yellow | amber | amber |
|---|---|---|---|---|---|---|---|---|
| 1 | User Identifier (first name, surname) | - | - | - | - | - | - | - |
| 2 | Date of filling the questionnaire | - | - | - | - | - | - | - |
| 3 | In which country do you currently work? | - | - | - | - | - | - | - |
| 4 | At which facility / study site do you currently work? | - | - | - | - | - | - | - |
| 5 | Which test are you assessing? | - | - | - | - | - | - | - |
| 6 | About how many times did you perform this test approximately? | - | - | - | - | - | - | - |
| 7 | About how many times did you observe the use of this test (not performed yourself)? | - | - | - | - | - | - | - |
| 8 | What is your profession? | - | - | - | - | - | - | - |
| 9 | How many years of laboratory experience do you have? | - | - | - | - | - | - | - |
| 10 | How many years of working experience in limited resource settings do you have? | - | - | - | - | - | - | - |
| 11 | How much experience do you have with interpreting the results of lateral flow tests or rapid diagnostics (e.g. for HIV, malaria, pregnancy)? | - | - | - | - | - | - | - |
| 12 | How satisfied were you with the following components of the test training? [Instructions for use] | very satisfied | satisfied | - | neither | - | dissatisfied | very dissatisfied |
| 12 | How satisfied were you with the following components of the test training? [Standard operating procedures] | - | - | - | - | - | - | - |
| 12 | How satisfied were you with the following components of the test training? [Face - to - face demonstration] | - | - | - | - | - | - | - |
| 13 | What additional materials (if any) do you think should be provided as part of the training? | None | - | - | 1-2 | - | >2 | - |
| 14 | How long should the training of this test be? | Self - explanatory, | 1 - 2 hours of train | 2 - 4 hours | Half a day | - | Full Day | - |
| 15 | Do you consider proficiency testing necessary? | No | - | - | - | - | Yes | - |
| 16 | Please comment here on the need for proficiency testing: | - | - | - | - | - | - | - |
| 17 | Do you think that company training is necessary? | No | - | - | - | - | Yes | - |
| 18 | Please comment on having the company come to do an in-person training. | - | - | - | - | - | - | - |
| 19 | After how many of these tests do you feel you could perform the test on your own? | 1 - 2 tests | - | - | 3 - 5 tests | - | 6 - 10 tests | 10 tests |
| 20 | How satisfied are you with the quality of each of the components in the kit (in terms of ease of use and fit for purpose)? [External paper box of the kit] | very satisfied | satisfied | - | neither | - | dissatisfied | very dissatisfied |
| 20 | How satisfied are you with the quality of each of the components in the kit (in terms of ease of use and fit for purpose)? [Extraction tube] | very satisfied | satisfied | - | neither | - | dissatisfied | very dissatisfied |
| 20 | How satisfied are you with the quality of each of the components in the kit (in terms of ease of use and fit for purpose)? [Filter cap] | very satisfied | satisfied | - | neither | - | dissatisfied | very dissatisfied |
| 20 | How satisfied are you with the quality of each of the components in the kit (in terms of ease of use and fit for purpose)? [Test cartridge / device] | very satisfied | satisfied | - | neither | - | dissatisfied | very dissatisfied |
| 20 | How satisfied are you with the quality of each of the components in the kit (in terms of ease of use and fit for purpose)? [Test cartridge - packing pouch] | very satisfied | satisfied | - | neither | - | dissatisfied | very dissatisfied |
| 20 | How satisfied are you with the quality of each of the components in the kit (in terms of ease of use and fit for purpose)? [Swab for specimen collection] | very satisfied | satisfied | - | neither | - | dissatisfied | very dissatisfied |
| 21 | Which kit component(s) should be improved in your opinion (if any)? | - | - | - | - | - | - | - |
| 22 | Overall, how satisfied are you with the kit components? | 1 | 2 | - | 3 | - | 4 | 5 |
| 23 | How satisfied are you with the design of the following features? [Size of the cartridge] | very satisfied | satisfied | - | neither | - | dissatisfied | very dissatisfied |
| 23 | How satisfied are you with the design of the following features? [Size of the well to add sample mix] | very satisfied | satisfied | - | neither | - | dissatisfied | very dissatisfied |
| 24 | How satisfied are you with the quality of the quality controls in terms of ease of use and fit for purpose | 1 | 2 | - | 3 | - | 4 | 5 |
| 25 | How useful do you find the inclusion of a positive control? | 1 | 2 | - | 3 | - | 4 | 5 |
| 26 | How useful do you find the inclusion of a negative control? | 1 | 2 | - | 3 | - | 4 | 5 |
| 27 | Please comment on the added value and quality of the quality controls. | - | - | - | - | - | - | - |
| 28 | How satisfied are you with the quality of the reader in terms of its ease of use and fit for purpose? | 1 | 2 | - | 3 | - | 4 | 5 |
| 29 | Overall, how satisfied are you with the Lumira instrument? | 1 | 2 | - | 3 | - | 4 | 5 |
| 30 | Which component(s) of the Lumira instrument should be improved in your opinion (if any) and why? | - | - | - | - | - | - | - |
| 31 | Please determine the difficulty of the following steps: [a) Check expiry date] | Very easy | Easy | - | Neither | - | Difficult | Very difficult |
| 31 | Please determine the difficulty of the following steps: [b) Label the assay diluent tube with the patient identifier] | Very easy | Easy | - | Neither | - | Difficult | Very difficult |
| 31 | Please determine the difficulty of the following steps: [c) Open the extraction tube by removing the seal (if appl.)] | Very easy | Easy | - | Neither | - | Difficult | Very difficult |
| 31 | Please determine the difficulty of the following steps: [d) Insert the swab into the tube] | Very easy | Easy | - | Neither | - | Difficult | Very difficult |
| 31 | Please determine the difficulty of the following steps: [e) Ease of swab extraction procedure] | Very easy | Easy | - | Neither | - | Difficult | Very difficult |
| 31 | Please determine the difficulty of the following steps: [f) Ability to perform extraction procedure consistently] | Very easy | Easy | - | Neither | - | Difficult | Very difficult |
| 31 | Please determine the difficulty of the following steps: [g) Ability to maintain cleanliness of ancillary devices (e.g. pipette) in order to avoid cross-contamination] | Very easy | Easy | - | Neither | - | Difficult | Very difficult |
| 31 | Please determine the difficulty of the following steps: [h) Remove the test cartridge from the pouch] | Very easy | Easy | - | Neither | - | Difficult | Very difficult |
| 31 | Please determine the difficulty of the following steps: [i) Ease of transferring sample onto device] | Very easy | Easy | - | Neither | - | Difficult | Very difficult |

| | | | | | | | | |
|---|---|---|---|---|---|---|---|---|
| 31 | Please determine the difficulty of the following steps: [i) Ease of transferring sample onto device] | Very easy | Easy | - | Neither | - | Difficult | Very difficult |
| 31 | Please determine the difficulty of the following steps: [j) Ease of transferring exact quantity into the sample well] | Very easy | Easy | - | Neither | - | Difficult | Very difficult |
| 31 | Please determine the difficulty of the following steps: [k) Ease of using the LumiraDx instrument] | Very easy | Easy | - | Neither | - | Difficult | Very difficult |
| 31 | Please determine the difficulty of the following steps: [l) Trouble shooting] | Very easy | Easy | - | Neither | - | Difficult | Very difficult |
| 32 | How satisfied are you with the logical sequence of steps? | 1 | 2 | - | 3 | - | 4 | 5 |
| 33 | Overall, how difficult did you find the steps? | 1 | 2 | - | 3 | - | 4 | 5 |
| 34 | Overall how satisfied are you with the time-relevant components (e.g. are the steps relatively long, are there many specific timed steps that make it hard to keep | 1 | 2 | - | 3 | - | 4 | 5 |
| 35 | Please assess the time relevant components of the test. [Pre-analytic time (i.e. from when you get the swab to when you start incubation)] | 2 min | - | - | 3 to 5 min | 6 to 10 min | 10 min | - |
| 35 | Please assess the time relevant components of the test. [Incubation time (i.e. how long do you have to wait to get the results once you add your sample to the stri | 15 min | 15 to 30 min | - | 30 to 60 min | - | 60 min | - |
| 35 | Please assess the time relevant components of the test. [Analytic time (i.e. time needed to analyse the results once the test conduction is completed)] | 2 min | - | - | 3 to 5 min | 6 to 10 min | 10 min | - |
| 36 | In your opinion, about how many patients could be tested with this test in an 8-hour day? | 100 | - | - | 50 - 100 | - | 10 - 50 | 10 |
| 37 | Please comment here if you see any potential issues or room for improvement. | - | - | - | - | - | - | - |
| 38 | How did you find the results read-out in the following areas: [a) Read-out from Reader] | Very easy | Easy | - | Neither | - | Difficult | Very difficult |
| 38 | How did you find the results read-out in the following areas: [b) Interpretation of the test result] | Very easy | Easy | - | Neither | - | Difficult | Very difficult |
| 39 | Do you foresee any issues with reading these results considering the lighting conditions in the settings you currently work or have experience with? | No | - | - | - | - | - | Yes (please explain below) |
| 39 | If yes, please explain here | - | - | - | - | - | - | - |
| 40 | Overall, how did you find the use of this rapid COVID-19 diagnostic tool? | 1 | 2 | - | 3 | - | 4 | 5 |
| 41 | Please comment here on the use: | - | - | - | - | - | - | - |
| 42 | Which option(s) do you consider feasible in your setting (assuming you only have one instrument and one sampling method)? | Sequential testing | - | - | - | - | Sequential testing | - |
| 43 | Which aspect(s) of this test could cause difficulties in its day-to-day use? | 0 | - | - | 1 | 2 | 3 | 4 |
| 44 | Please give a short explanation for each of the aspects you selected above e.g. what could be the challenges in the day-to-day use: | - | - | - | - | - | - | - |
| 45 | Do you see this test being used in its current form in your setting in your country? | Yes | | - | - | - | No | - |
| 45 | Please elaborate: | - | - | - | - | - | - | - |
| 46 | If yes, at which health care level(s) do you see this test being implemented in your country? | At a testing site o | Family doctor / Ge | - | Peripheral hospita | - | Reference hospita | None |
| 47 | If you don't see this test being used in its current form, which aspects should be changed to make it suitable for use in your setting in your country: | none | - | - | 1-2 | - | >2 | - |
| 48 | Do you see this test being used in its current form in your setting in low and middle income countries? | Yes (please expla | - | - | - | - | No (please explai | - |
| 48 | Please elaborate: | - | - | - | - | - | - | - |
| 49 | At which health care level(s) do you see this test being implemented in low and middle income countries? | Primary health car | Famildy doctor / G | Health centre / mi | District hospital / | - | Reference hospita | None |
| 50 | If you don't see this test being used in its current form, which aspects should be changed to make it suitable for use in your setting in low and middle income c | none | - | - | 1 | 2 | 3 | 4 |
| 52 | Anything else you would like to add? | - | - | - | - | - | - | - |

#### (E) Table 1: Detailed list of symptoms for all PCR positives

| Viral load *(log₁₀RNA copies/mL)* | Result Ag-RDT | Increased temperature/ fever | Cough | Do you have a productive cough? | Sore throat | Shortness of breath | Muscle pain/Body aches | Fatigue | Headache | Runny nose | Chest pain | Diarrhea | Nausea/ vomiting | Loss of taste /smell | Other symptoms |
|---|---|---|---|---|---|---|---|---|---|---|---|---|---|---|---|
| 6.85 | positive | No | Yes | Yes | No | No | No | Yes | Yes | Yes | No | No | No | Yes | |
| 7.56 | positive | No | Yes | No | Yes | No | Yes | Yes | Yes | Yes | No | No | No | Yes | No |
| 7.97 | positive | Yes | Yes | No | Yes | No | Yes | Yes | Yes | Yes | No | No | No | Yes | Yes |
| 7.76 | positive | No | No | No | Yes | Yes | Yes | Yes | Yes | No | Yes | No | No | No | No |
| 8.00 | positive | Yes | Yes | No | No | No | Yes | Yes | Yes | No | Yes | No | No | No | Yes |
| 7.61 | positive | Yes | Yes | Yes | No | No | Yes | Yes | No | No | No | No | No | No | No |
| 7.11 | negative | No | No | No | No | No | No | No | No | No | No | No | No | Yes | No |
| 8.15 | positive | Yes | Yes | No | Yes | No | Yes | No | Yes | Yes | No | No | No | Yes | No |
| 6.82 | positive | | | | | | | | | | | | | | |
| 5.99 | positive | Yes | No | No | No | No | Yes | Yes | No | No | No | No | No | No | No |
| 7.79 | positive | No | Yes | No | Yes | No | Yes | No | Yes | No | No | No | No | No | No |
| 3.99 | positive | No | No | No | No | Yes | No | Yes | Yes | No | No | No | No | No | No |
| 6.32 | positive | No | Yes | Yes | No | No | Yes | Yes | Yes | Yes | No | No | No | No | No |
| 8.38 | positive | No | Yes | | Yes | No | Yes | Yes | Yes | Yes | No | No | No | No | No |
| 4.26 | negative | No | No | No | No | No | Yes | Yes | No | No | No | No | No | No | Yes |
| 7.73 | positive | No | No | No | No | No | Yes | Yes | Yes | No | No | No | No | Yes | Yes |
| 8.48 | positive | No | Yes | No | Yes | No | No | Yes | Yes | Yes | No | No | No | Yes | No |
| 8.20 | positive | Yes | Yes | No | No | Yes | Yes | Yes | Yes | Yes | No | Yes | No | Yes | No |
| 8.48 | positive | No | Yes | No | No | Yes | Yes | Yes | No | Yes | No | No | No | Yes | No |
| 8.32 | positive | Yes | Yes | Yes | No | No | Yes | No | Yes | Yes | No | No | No | No | No |
| 6.18 | positive | Yes | No | No | No | No | No | Yes | No | Yes | No | No | No | Yes | Yes |
| 3.78 | negative | No | Yes | No | Yes | No | No | No | No | Yes | No | No | No | Yes | No |
| 7.61 | positive | | | | | | | | | | | | | | |
| 5.56 | positive | | Yes | No | No | No | Yes | Yes | Yes | Yes | No | No | No | Yes | No |
| 3.99 | positive | | | | | | | | | | | | | | |
| 7.73 | positive | Yes | Yes | No | Yes | No | Yes | Yes | Yes | No | Yes | Yes | No | No | No |
| 8.08 | positive | No | No | No | No | No | Yes | Yes | Yes | No | No | Yes | No | No | No |

| | | | | | | | | | | | | | | | |
|---|---|---|---|---|---|---|---|---|---|---|---|---|---|---|---|
| 7.53 | positive | Yes | Yes | No | Yes | No | Yes | Yes | Yes | Yes | No | Yes | No | Yes | No |
| 7.56 | positive | No | Yes | No | Yes | No | No | No | Yes | Yes | No | No | No | No | No |
| 7.83 | positive | Yes | No | No | No | No | Yes | Yes | Yes | No | No | No | No | No | No |
| 6.67 | positive | | | | | | | | | | | | | | |
| 6.58 | positive | No | Yes | No | No | No | Yes | Yes | Yes | No | Yes | Yes | No | No | No |
| 4.90 | positive | Yes | No | No | Yes | No | No | No | Yes | No | No | No | No | No | No |
| 5.40 | positive | No | Yes | No | Yes | No | No | Yes | Yes | No | No | No | Yes | No | No |
| 2.95 | negative | | | | | | | | | | | | | | |
| 8.59 | positive | Yes | Yes | Yes | No | No | Yes | No | No | Yes | No | No | No | No | No |
| 6.04 | positive | Yes | | | | | | | | | | | | | |
| 4.81 | negative | No | Yes | No | No | No | No | Yes | Yes | No | No | No | No | Yes | |
| 8.08 | positive | No | Yes | No | No | No | No | Yes | No | No | No | No | No | No | No |
| 7.41 | positive | Yes | No | No | No | No | Yes | Yes | Yes | No | No | Yes | No | No | No |
| 8.00 | positive | No | No | No | Yes | No | No | No | No | Yes | No | No | No | No | No |
| 4.79 | positive | Yes | Yes | Yes | No | No | Yes | Yes | Yes | No | No | Yes | No | No | No |
| 6.94 | positive | Yes | No | Yes | No | No | Yes | Yes | No | Yes | No | No | No | Yes | |
| 5.52 | positive | No | No | No | No | No | Yes | Yes | Yes | No | No | Yes | No | No | No |
| 7.04 | positive | No | No | No | Yes | No | Yes | Yes | Yes | No | No | No | No | Yes | No |
| 6.28 | positive | | | | | | | | | | | | | | |
| 8.11 | positive | No | Yes | Yes | No | No | No | Yes | No | Yes | No | No | No | Yes | No |
| 5.11 | positive | Yes | Yes | Yes | Yes | No | Yes | Yes | Yes | Yes | Yes | No | No | Yes | No |
| 7.71 | positive | Yes | Yes | No | Yes | No | Yes | Yes | Yes | Yes | No | No | No | No | No |
| 3.66 | negative | Yes | Yes | No | Yes | No | No | Yes | Yes | No | No | Yes | No | Yes | Yes |
| 6.82 | positive | | | | | | | | | | | | | | |
| 7.15 | positive | No | No | No | No | No | No | No | No | Yes | No | No | No | No | No |
| 7.85 | positive | No | No | No | Yes | Yes | Yes | Yes | Yes | Yes | Yes | No | No | No | No |
| 6.97 | positive | No | Yes | No | Yes | No | No | Yes | Yes | No | Yes | No | No | Yes | No |
| 2.60 | negative | No | Yes | No | Yes | Yes | No | Yes | Yes | No | No | No | No | No | Yes |
| 4.08 | negative | | | | | | | | | | | | | | |
| 8.80 | positive | No | Yes | | Yes | | Yes | Yes | Yes | Yes | No | No | No | No | No |
| 6.61 | negative | No | Yes | No | No | No | No | Yes | Yes | No | No | No | No | No | Yes |
| 4.36 | positive | No | No | No | Yes | No | No | Yes | No | No | No | No | No | Yes | No |
| 4.18 | negative | No | No | No | No | Yes | Yes | Yes | Yes | Yes | Yes | No | No | Yes | Yes |
| 5.38 | positive | | | | | | | | | | | | | | |

| | | | | | | | | | | | | | | | |
|---|---|---|---|---|---|---|---|---|---|---|---|---|---|---|---|
| 6.04 | positive | No | No | No | No | No | No | No | No | Yes | No | No | No | No | No |
| 7.08 | positive | No | Yes | No | No | No | No | No | No | No | No | No | No | No | No |
| 7.45 | positive | Yes | Yes | Yes | No | No | No | Yes | Yes | No | No | No | No | No | No |
| 4.26 | positive | No | Yes | Yes | No | No | Yes | Yes | Yes | No | No | No | Yes | Yes | No |
| 7.79 | positive | No | Yes | Yes | No | No | No | No | Yes | No | Yes | No | No | No | No |
| 6.07 | negative | No | No | No | No | No | No | Yes | No | No | No | No | No | Yes | No |
| 8.25 | positive | No | Yes | Yes | No | No | Yes | | Yes | Yes | No | No | No | No | No |
| 7.07 | positive | No | Yes | No | No | No | Yes | Yes | No | Yes | No | No | Yes | No | No |
| 9.34 | positive | No | Yes | Yes | No | No | Yes | No | No | Yes | No | No | No | No | No |
| 9.08 | positive | No | Yes | No | No | No | No | Yes | No | Yes | No | Yes | No | No | No |
| 7.28 | positive | No | No | No | Yes | No | Yes | Yes | Yes | Yes | No | No | No | Yes | No |
| 7.53 | positive | No | Yes | No | Yes | No | Yes | Yes | Yes | Yes | No | No | Yes | | No |
| 5.47 | positive | No | Yes | No | Yes | No | No | No | No | Yes | No | No | No | No | No |
| 9.03 | positive | Yes | No | No | No | No | No | Yes | No | No | No | No | | No | No |
| 8.99 | positive | Yes | Yes | No | No | No | Yes | Yes | No | Yes | No | No | No | No | No |
| 8.61 | positive | No | Yes | No | Yes | No | Yes | No | No | No | No | No | No | No | No |
| 8.12 | positive | No | No | No | Yes | Yes | No | No | Yes | No | No | No | No | Yes | No |
| 9.37 | positive | No | Yes | No | No | No | No | No | Yes | Yes | No | No | No | No | Yes |
| 6.58 | positive | No | Yes | No | Yes | No | Yes | Yes | No | No | No | No | No | Yes | No |
| 6.19 | positive | No | No | No | No | No | Yes | Yes | Yes | Yes | Yes | No | Yes | Yes | No |
| 6.93 | positive | No | No | No | No | No | No | Yes | No | Yes | No | No | No | Yes | No |
| 4.94 | negative | No | No | No | Yes | No | No | No | No | No | No | No | No | No | No |
| 9.43 | positive | Yes | Yes | No | Yes | No | Yes | Yes | Yes | No | No | No | No | No | No |
| 5.04 | negative | No | Yes | No | No | No | No | Yes | No | No | No | No | No | Yes | No |
| 9.30 | negative | No | No | No | Yes | No | No | Yes | No | No | No | No | No | No | No |
| 8.77 | positive | Yes | Yes | No | No | Yes | Yes | Yes | No | Yes | No | No | No | No | No |
| 8.10 | positive | No | Yes | No | No | No | No | No | No | Yes | No | No | No | No | No |
| 9.29 | positive | No | Yes | No | No | No | No | Yes | No | No | No | No | No | No | No |
| 6.97 | negative | No | No | No | No | No | No | No | No | No | No | No | No | Yes | No |
| 4.94 | positive | No | No | No | No | No | No | Yes | No | Yes | No | No | No | Yes | No |
| 8.14 | positive | Yes | Yes | Yes | Yes | No | Yes | Yes | Yes | Yes | No | Yes | No | Yes | No |
| 5.61 | positive | Yes | Yes | No | No | No | No | Yes | No | No | No | No | No | No | No |
| 6.94 | positive | No | No | No | No | No | No | No | No | Yes | No | No | No | Yes | No |
| 7.07 | positive | No | No | No | No | No | No | No | No | No | No | No | No | Yes | No |

| | | | | | | | | | | | | | | | |
|---|---|---|---|---|---|---|---|---|---|---|---|---|---|---|---|
| 5.76 | positive | No | No | No | No | No | No | No | No | No | No | No | No | Yes | No |
| 5.68 | positive | No | No | No | No | No | No | Yes | No | No | No | No | No | Yes | No |
| 8.56 | positive | Yes | No | | No | No | No | Yes | No | No | No | No | No | No | No |
| 5.42 | negative | No | No | No | No | No | No | No | No | No | No | No | No | Yes | No |
| 6.80 | positive | No | Yes | No | No | No | No | Yes | No | No | No | No | No | Yes | No |
| 5.95 | positive | No | No | No | No | No | Yes | Yes | No | Yes | No | No | No | Yes | No |
| 3.72 | negative | No | Yes | No | No | No | No | No | Yes | No | No | No | No | Yes | No |
| 3.57 | negative | No | No | No | No | No | No | No | No | No | No | No | No | Yes | No |
| 6.23 | negative | No | Yes | No | No | No | No | Yes | No | Yes | No | No | No | Yes | No |
| 6.87 | positive | No | No | No | No | No | No | No | Yes | No | No | No | No | Yes | No |
| 8.08 | negative | No | Yes | No | No | No | Yes | Yes | No | No | No | No | No | No | No |
| 6.95 | positive | No | No | No | Yes | No | No | No | No | No | No | No | No | No | No |
| 5.88 | positive | No | Yes | Yes | No | No | No | No | No | No | No | No | No | Yes | No |
| 6.81 | positive | No | No | No | No | No | No | No | No | Yes | No | No | No | No | No |
| 8.65 | positive | No | No | No | No | No | Yes | Yes | No | Yes | No | No | No | No | No |
| 6.83 | positive | No | No | No | Yes | No | No | Yes | Yes | No | No | No | No | Yes | No |
| 8.64 | positive | No | Yes | No | Yes | No | Yes | Yes | Yes | No | No | No | No | Yes | No |
| 7.85 | positive | No | Yes | Yes | Yes | No | Yes | Yes | Yes | Yes | No | Yes | No | No | No |
| 8.32 | positive | No | No | No | Yes | No | Yes | Yes | Yes | No | No | No | | No | No |
| 9.27 | negative | No | Yes | | Yes | No | No | Yes | Yes | No | No | No | | No | No |
| 8.89 | positive | Yes | Yes | Yes | Yes | No | Yes | No | Yes | Yes | No | Yes | No | Yes | No |
| 4.46 | positive | Yes | No | No | Yes | No | No | Yes | Yes | No | No | Yes | No | Yes | No |
| 6.58 | positive | No | No | No | Yes | No | No | No | No | No | No | No | No | Yes | No |
| 7.75 | negative | No | Yes | No | No | No | Yes | Yes | Yes | Yes | No | No | No | No | No |
| 9.69 | positive | Yes | No | No | No | No | Yes | Yes | No | No | No | No | No | Yes | |
| 8.91 | positive | No | Yes | No | Yes | No | Yes | Yes | Yes | No | No | Yes | No | Yes | No |
| 8.70 | positive | Yes | Yes | Yes | No | No | No | No | Yes | No | No | No | No | No | No |
| 8.17 | positive | No | Yes | | No | No | No | Yes | No | No | No | No | No | No | No |
| 8.72 | positive | No | Yes | No | No | No | No | No | No | No | No | No | No | Yes | No |
| 3.83 | negative | No | No | No | Yes | No | No | No | No | No | No | No | No | No | No |
| 7.86 | positive | No | Yes | No | Yes | No | Yes | Yes | Yes | Yes | No | No | No | Yes | No |
| 8.75 | positive | No | No | No | No | No | Yes | No | No | No | No | No | No | No | No |
| 7.82 | positive | No | Yes | Yes | No | No | No | Yes | Yes | No | No | Yes | No | No | No |
| 8.01 | positive | No | No | No | Yes | No | No | No | No | Yes | No | No | No | Yes | No |

| | | | | | | | | | | | | | | | |
|---|---|---|---|---|---|---|---|---|---|---|---|---|---|---|---|
| 9.08 | positive | No | Yes | No | No | No | Yes | No | Yes | No | No | No | No | No | Yes |
| 9.63 | positive | No | No | No | No | No | No | Yes | No | Yes | No | No | No | No | No |
| 7.47 | positive | No | Yes | Yes | No | No | Yes | Yes | Yes | Yes | No | Yes | Yes | No | No |
| 7.24 | positive | No | No | No | No | No | Yes | No | Yes | Yes | No | No | No | Yes | No |
| 8.67 | positive | Yes | Yes | No | No | No | Yes | Yes | Yes | Yes | No | No | No | Yes | No |
| 8.47 | positive | Yes | No | Yes | No | No | Yes | Yes | Yes | No | No | No | No | No | No |
| 6.03 | negative | No | No | No | No | No | No | Yes | Yes | No | No | No | No | Yes | No |
| 5.86 | negative | Yes | No | No | No | No | Yes | Yes | Yes | No | No | No | No | Yes | No |
| 6.81 | positive | No | Yes | Yes | Yes | No | Yes | Yes | Yes | Yes | No | No | No | Yes | No |
| 4.70 | negative | No | Yes | No | No | No | No | No | No | No | No | No | | Yes | |
| 6.61 | positive | No | Yes | | No | No | No | No | No | Yes | No | No | | Yes | No |
| 7.34 | positive | | | | | | Yes | | | | | | | Yes | |
| 8.26 | positive | No | Yes | Yes | Yes | No | No | Yes | No | No | No | No | No | Yes | No |
| 5.61 | positive | No | Yes | No | No | No | No | No | Yes | No | No | No | No | Yes | No |
| 9.00 | positive | Yes | No | No | No | No | No | Yes | No | Yes | No | No | No | Yes | No |
| 8.38 | positive | Yes | No | No | No | No | No | No | No | No | No | No | No | No | Yes |
| 9.19 | positive | No | No | No | Yes | No | Yes | Yes | Yes | No | Yes | No | No | Yes | No |

#### (F) Table 2: Antigen-based RDT with test result, CT values and viral load for PCR positive participants in Berlin and Heidelberg

CT-value reported here represent the E-gene genome target (similar to the target described by Corman) in descending order. A conversion of CT-values for RT-PCR tests into viral-load was performed using quantified specific in vitro-transcribed RNA (Corman 2020 Eurosurveillance).

| Berlin | | | |
|---|---|---|---|
| **Antigen RDT result** | **CT-value** (E-Gene) | **Viral load** ($log_{10}$ SARS-CoV2 RNA copies /mL) | **PCR assay** |
| positive | 14.61 | 9.63 | TIB Molbiol |
| positive | 15.29 | 9.43 | TIB Molbiol |
| positive | 15.51 | 9.37 | TIB Molbiol |
| positive | 15.60 | 9.34 | TIB Molbiol |
| negative | 15.74 | 9.30 | TIB Molbiol |
| positive | 15.76 | 9.29 | TIB Molbiol |
| negative | 15.84 | 9.27 | TIB Molbiol |
| positive | 16.09 | 9.19 | TIB Molbiol |
| positive | 16.46 | 9.08 | TIB Molbiol |
| positive | 16.64 | 9.03 | TIB Molbiol |
| positive | 16.76 | 9.00 | TIB Molbiol |

| | | | |
|---|---|---|---|
| positive | 17.12 | 8.89 | TIB Molbiol |
| positive | 17.53 | 8.77 | TIB Molbiol |
| positive | 17.60 | 8.75 | TIB Molbiol |
| positive | 17.69 | 8.72 | TIB Molbiol |
| positive | 17.75 | 8.70 | TIB Molbiol |
| positive | 17.85 | 8.67 | TIB Molbiol |
| positive | 17.93 | 8.65 | TIB Molbiol |
| positive | 18.04 | 8.61 | TIB Molbiol |
| positive | 18.82 | 8.38 | TIB Molbiol |
| positive | 19.04 | 8.32 | TIB Molbiol |
| positive | 19.23 | 8.26 | TIB Molbiol |
| positive | 19.27 | 8.25 | TIB Molbiol |
| positive | 19.63 | 8.14 | TIB Molbiol |
| positive | 19.70 | 8.12 | TIB Molbiol |
| positive | 19.77 | 8.10 | TIB Molbiol |
| negative | 19.85 | 8.08 | TIB Molbiol |
| positive | 20.08 | 8.01 | TIB Molbiol |
| positive | 20.58 | 7.86 | TIB Molbiol |
| positive | 21.88 | 7.47 | TIB Molbiol |
| positive | 22.34 | 7.34 | TIB Molbiol |

| | | | |
|---|---|---|---|
| positive | 22.52 | 7.28 | TIB Molbiol |
| positive | 22.66 | 7.24 | TIB Molbiol |
| positive | 23.25 | 7.07 | TIB Molbiol |
| negative | 23.57 | 6.97 | TIB Molbiol |
| positive | 23.65 | 6.95 | TIB Molbiol |
| positive | 23.69 | 6.94 | TIB Molbiol |
| positive | 23.91 | 6.87 | TIB Molbiol |
| positive | 24.10 | 6.81 | TIB Molbiol |
| positive | 24.11 | 6.81 | TIB Molbiol |
| positive | 24.15 | 6.80 | TIB Molbiol |
| positive | 24.78 | 6.61 | TIB Molbiol |
| negative | 26.07 | 6.23 | TIB Molbiol |
| negative | 26.61 | 6.07 | TIB Molbiol |
| positive | 26.99 | 5.95 | TIB Molbiol |
| positive | 27.23 | 5.88 | TIB Molbiol |
| negative | 27.30 | 5.86 | TIB Molbiol |
| positive | 28.14 | 5.61 | TIB Molbiol |
| positive | 28.16 | 5.61 | TIB Molbiol |
| positive | 28.62 | 5.47 | TIB Molbiol |
| negative | 28.78 | 5.42 | TIB Molbiol |

| | | | |
|---|---|---|---|
| negative | 31.20 | 4.70 | TIB Molbiol |
| negative | 34.14 | 3.83 | TIB Molbiol |
| negative | 34.50 | 3.72 | TIB Molbiol |
| negative | 35.00 | 3.57 | TIB Molbiol |

**(G) Figure 2: Correlation between the cut-off-index value and the viral load for all RT-PCR positive cases**

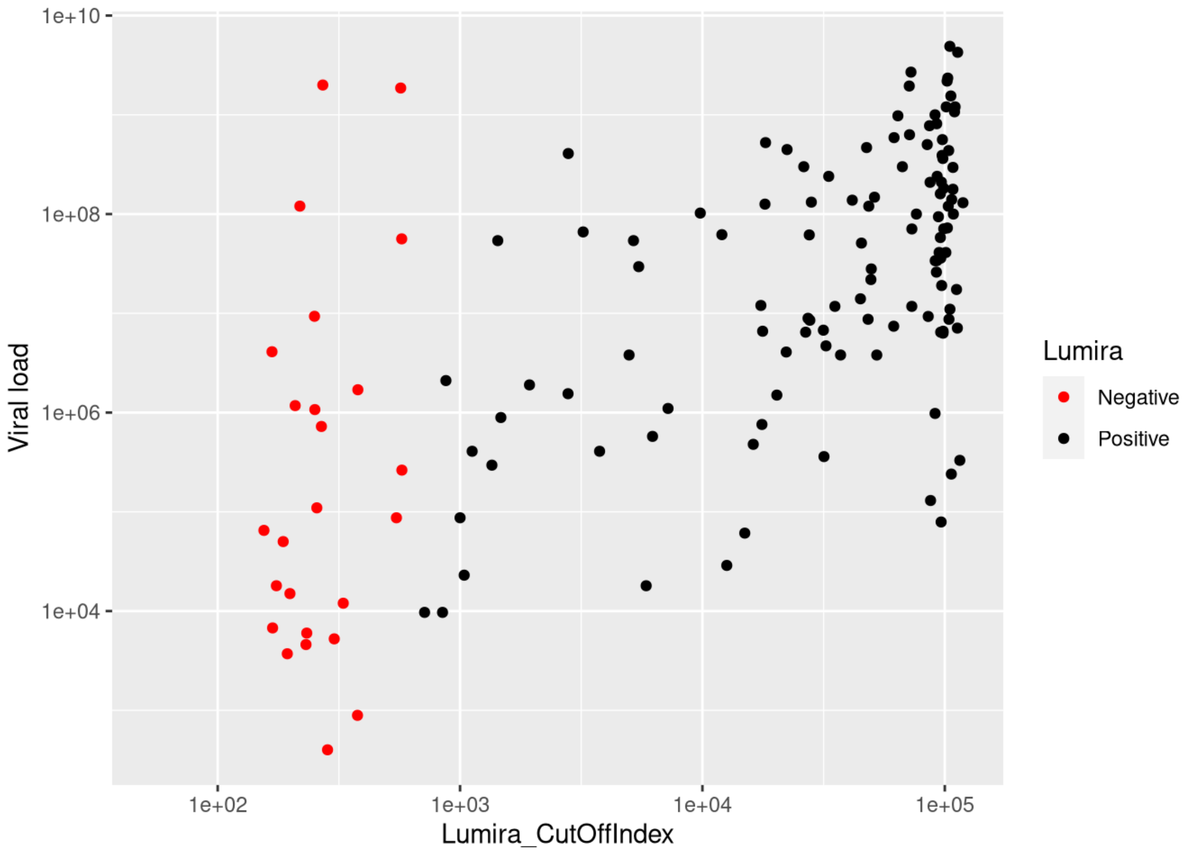

#### (H) Table 3: Discrepant analysis

| PCR | Ag-Test Lumira | COI | CT value: E-Gene | Viral load | Buffer PCR | Buffer CT value: E-Gene | Buffer Viral load |
|---|---|---|---|---|---|---|---|
| positive | negative | | 22,47 | 7,11 | | | |
| positive | negative | | 32,18 | 4,26 | | | |
| positive | negative | | 33,83 | 3,78 | | | |
| positive | negative | | 36,61 | 2,95 | negative | | |
| positive | negative | | 30,25 | 4,81 | negative | | |
| positive | negative | | 34,16 | 3,66 | negative | | |
| positive | negative | 284 | 37,82 | 2,6 | negative | | |
| positive | negative | 330 | 32,75 | 4,08 | negative | | |
| positive | negative | 168 | 24,21 | 6,61 | negative | | |
| positive | negative | 199 | 32,46 | 4,18 | negative | | |
| positive | negative | | 26,61 | 6,07 | | | |
| positive | negative | 546 | 33,55 | 4,94 | | | |
| positive | negative | | 33,21 | 5,04 | | | |
| positive | negative | | 15,74 | 9,3 | positive | 34,9 | 4,52 |
| positive | negative | 251 | 23,57 | 6,97 | | | |
| positive | negative | 575 | 28,78 | 5,42 | | | |
| positive | negative | 303 | 34,5 | 3,72 | | | |
| positive | negative | 194 | 35 | 3,57 | | | |
| positive | negative | 379 | 26,07 | 6,23 | | | |
| positive | negative | 218 | 19,85 | 8,08 | | | |
| positive | negative | | 15,84 | 9,27 | positive | | < 100 CP/mL |
| positive | negative | | 24,05 | 7,75 | positive | 37,82 | 2,61 |
| positive | negative | 168 | 34,14 | 3,83 | | | |
| positive | negative | 252 | 29,87 | 6,03 | | | |
| positive | negative | 268 | 27,03 | 5,86 | | | |
| positive | negative | 186 | 31,2 | 4,7 | | | |

| | | | | | |
|---|---|---|---|---|---|
| negative | positive | | | | negative |
| negative | positive | | | | negative |
| negative | positive | | | | negative |
| negative | positive | 687 | | | |

#### (I) Table 4: Exclusivity testing of LumiraDx™

| Pathogen | CT (lowest - highest value) *concentration (colony forming units [CFU] /ml)* | LumiraDx™ | Cross-Reactivity |
|---|---|---|---|
| Coronavirus OC 43<br>Coronavirus NL 63<br>Coronavirus 229 E<br>Coronavirus HKU 1 | 19.8 - 27.7<br>25.9 - 30.6<br>22.7 - 27.0<br>18.1 - 31.5 | 3/3 negative<br>3/3 negative<br>3/3 negative<br>3/3 negative | no |
| Adenovirus | 26.7 - 35 | 5/5 negative | no |
| Bocavirus | 31.5 - 33.2 | 2/2 negative | no |
| Influenza A H3N2 virus<br>Influenza A H1N1 virus<br>Influenza B virus | 17.6 - 30.7<br>19.4 - 29.9<br>16.8 - 30.5 | 4/4 negative<br>3/3 negative<br>3/3 negative | no |
| Metapneumovirus | 26.2 - 32.7 | 4/4 negative | no |
| Parainfluenza virus | 17.4 - 32.5 | 10/10 negative | no |
| Respiratory syncytial virus | 16.4 - 31.2 | 10/10 negative | no |
| Rhinovirus | 28.5 - 35 | 10/10 negative | no |
| *Mycoplasma pneumoniae* | 23.2 - 35 | 8/8 negative | no |
| *Staphylococcus aureus* | Unknown (clinical samples) | 9/9 negative | no |
| *Streptococcus sp*. | $1x10^7$ KBE/ml | 7/7 negative | no |

#### (J) Figure 3: System Usability Score and Ease-of-Use assessment results

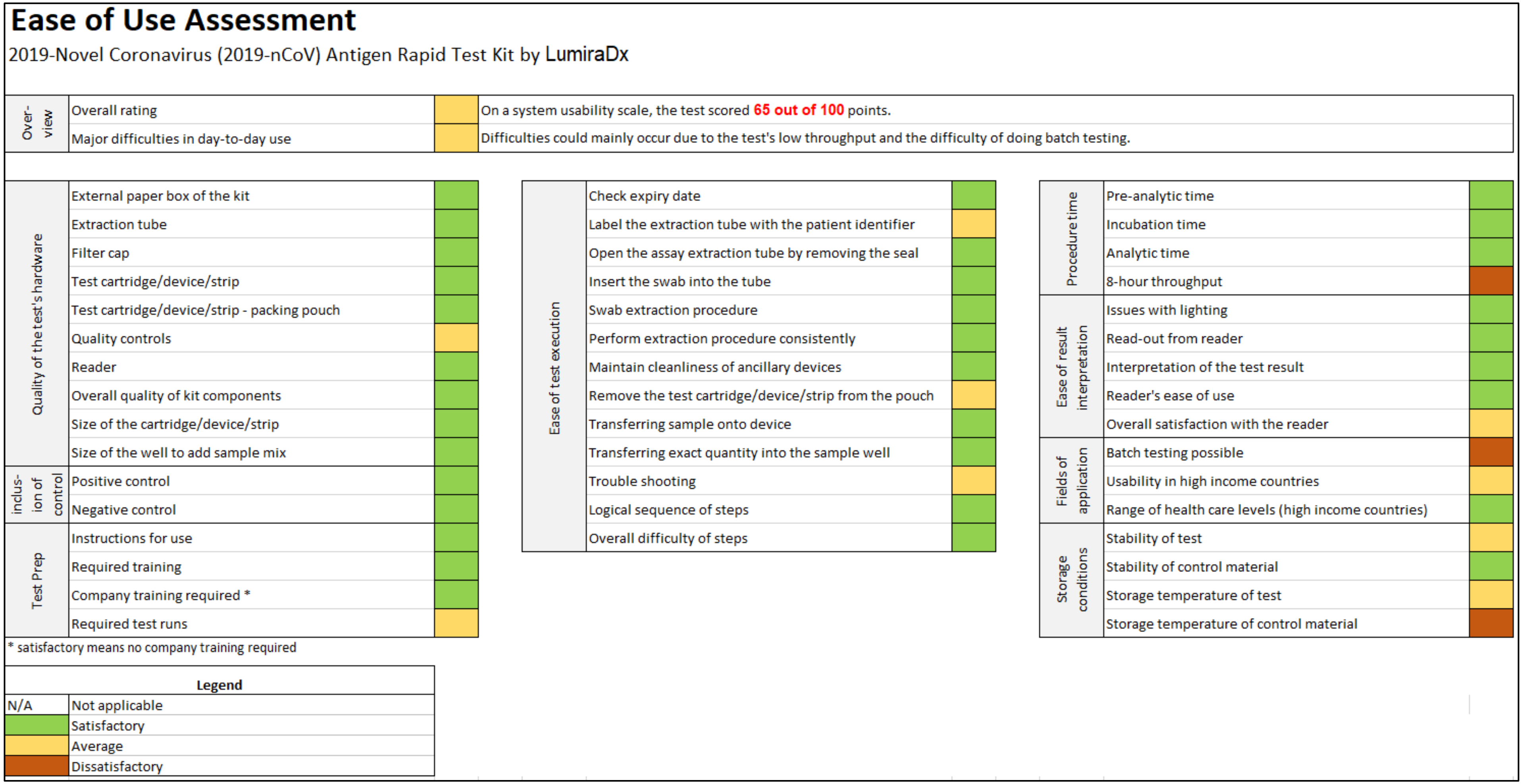

#### (K) Table 5: Comorbidities and list of symptoms of participants overall, Berlin and Heidelberg

| | Overall | Heidelberg | Berlin |
|---|---|---|---|
| **Data combined: Overweight and Adipositas > BMI 25** – Information available on N=703 | | | |
| Yes | 327 (46.5%) | 243 (50.2%) | 84 (38.4%) |
| No | 376 (53.5%) | 241 (49.8%) | 135 (61.6%) |
| **Comorbidities** – Information available on N=760 | | | |
| All with comorbidities | 241 | 183 | 58 |
| Lung diseases | | | |
| Asthma bronchiale | 60 | 39 | 21 |
| Chronic obstructive pulmonary disease (COPD) | 7 | 5 | 2 |
| Breathing disorders during sleep and obstructive Sleep Apnea (OSAS) | 9 | 9 | 0 |
| Interstitial Lung Disease | 0 | 0 | 0 |
| Lung Cancer | 2 | 1 | 1 |
| Other | 13 | 11 | 2 |
| Other diseases | | | |
| Cardiovascular diseases | 80 | 63 | 17 |
| Chronic kidney diseases | 9 | 8 | 1 |
| Autoimmune | 51 | 48 | 3 |
| HIV | 1 | 1 | 0 |
| Others | 79 | 52 | 27 |
| List of symptoms reported | | | |
| Fever | 90 (19.0%) | 49 (23.3%) | 41 (15.5%) |
| Fever measured<br>a. <38.4<br>b. >38.5 and <=39.4<br>c. >39.5 and >40.5 | <br>52(74.3%)<br>17(24.3%)<br>1 (1.4%) | <br>37(82.2%)<br>8(17.8%)<br>0 | <br>15(60%)<br>9(36%)<br>1(4%) |
| Cough | 247 (51.6%) | 112 (52.6%) | 135 (50.8%) |
| Productive cough | 72 (15.5%) | 40 (18.9%) | 32 (12.6%) |
| Sore throat | 242 (50.2%) | 115 (53.5%) | 127 (47.6%) |
| Shortness of breath | 46 (9.7%) | 31 (14.8%) | 15 (5.7%) |
| Muscle pain | 176 (36.7%) | 81 (38.2%) | 95 (35.4%) |
| Fatigue | 297 (62.0%) | 143 (66.8%) | 154 (58.1%) |
| Headache | 251 (52.3%) | 130 (60.7%) | 121 (45.5%) |
| Runny nose | 197 (41.5%) | 87 (41.6%) | 110 (41.4%) |
| Chest pain | 41 | 35 | 6 |

| | | | |
|---|---|---|---|
| | (8.6%) | (16.6%) | (2.3%) |
| Diarrhea | 54 (11.3%) | 25 (11.8%) | 29 (10.8%) |
| Nausea | 25 (5.4%) | 16 (4.3%) | 9 (6.3%) |
| Loss of taste and smell | 104 (21.8%) | 32 (15.3%) | 72 (27.0%) |

#### (L) Table 6: Sensitivity and Specificity overall and by subgroups

| | **Overall** N | **Ag-Test positive/ PCR positive** N (%) | **Ag-Test negative/ PCR positive** N (%) | **Ag-Test positive/ PCR negative** N (%) | **Ag-Test negative/ PCR negative** N (%) | **Sensitivity** % (95% CI) | **Specificity** % (95% CI) |
|---|---|---|---|---|---|---|---|
| **Sensitivity** | | | | | | | |
| **Overall** | 761 | 120 (15.8) | 26 (3.4) | 4 (0.5) | 611 (79.7) | 82.2 (75.2-87.5) | 99.3 (98.3-99.7) |
| **Heidelberg** | 488 | 55 (11.3) | 10 (2.0) | 3 (0.6) | 420 (86.1) | 84.6 (73.9-91.4) | 99.3 (97.9-99.8) |
| **Berlin** | 273 | 65 (23.8) | 16 (5.9) | 1 (0.4) | 191 (70.0) | 80.2 (70.3-87.5) | 99.5 (97.1-100) |
| **Symptom duration** – Information available for N=472 | | | | | | | |
| **0-7 days** Overall | 423 | 102 (24.1) | 16 (3.8) | 2 (0.5) | 303 (71.6) | 86.4 (79.1-91.5) | 99.3 (97.6-99.8) |
| **8-14 days** Overall | 39 | 7 (17.9) | 6 (15.4) | 0 | 26 (66.7) | 53.8 (29.1-76.8) | 100 (87.1-100) |
| **Symptomatic versus Asymptomatic** – Information available for N= 472 | | | | | | | |
| **Symptomatic** | 210 | 45 (21.4) | 8 (3.8) | 2 (1.0) | 155 (73.8) | 84.9 (72.9-92.1) | 98.7 (95.5-99.6) |
| **Asymptomatic** | 271 | 7 (2.6) | 2 (0.7) | 1 (0.4) | 261 (96.3) | 77.8 (45.3-93.7) | 99.6 (97.9-100) |
| **CT-Value PCR <30 and >=30** – Information available for N=146 | | | | | | | |
| **CT value PCR <30** | 122 | 110 | 12 | NA | NA | 90.2 (83.6-94.3) | NA |
| **CT value PCR ≥30** | 24 | 10 | 14 | NA | NA | 41.7 (24.5-61.2) | NA |
| **CT-Value PCR <25 and >=25** – Information available for N=146 | | | | | | | |
| **CT value PCR <25** | 95 | 88 | 7 | NA | NA | 92.6 (85.6-96.4) | NA |
| **CT value PCR ≥25** | 51 | 32 | 19 | NA | NA | 62.7 (49.0-74.7) | NA |
